# Multi-ancestry admixture mapping reveals ancestry-associated disease loci in the UK Biobank

**DOI:** 10.64898/2026.08.06.26359859

**Authors:** Riccardo Smeriglio, Sonia Moreno-Grau, Daniel Mas Montserrat, Guhan Venkataraman, David Bonet, Caterina Fuses, Manuel A. Rivas, Alessandro Savino, Stefano Di Carlo, Jordi Abante, Alexander G. Ioannidis

## Abstract

Genome-wide association studies have successfully identified thousands of genetic associations, yet their predominant reliance on European-descent populations limits insights into the full spectrum of human genetic diversity and its impact on disease. Admixture mapping offers a powerful, complementary approach by leveraging differences in haplotype frequencies across ancestral backgrounds to identify risk loci for complex traits. Here, we perform a large-scale, multi-ancestry admixture mapping study across 415,792 unrelated individuals in the UK Biobank, examining associations between local haplotype ancestry and 108 phenotypes. Our approach identifies 13 genome-wide significant ancestry-phenotype associations, recovering previously reported signals while uncovering four novel ancestry-associated findings, including new risk loci for atrial fibrillation, dermatitis, and angina pectoris. To overcome the limited resolution of traditional admixture mapping, we implemented a conditional fine-mapping framework, which enabled us to localize four putatively causal variants. *In silico* variant effect prediction and eQTL integration revealed regulatory and missense effects predominantly localized to lung, and immune tissues, aligning with captured phenotypes such as asthma, dermatitis, and hypothyroidism. Notably, our findings demonstrate striking genetic heterogeneity, revealing how the same clinical phenotype can arise through distinct genetic pathways depending on the ancestral background. Overall, this work highlights the critical importance of modeling local ancestry structure to refine genetic associations, uncover novel disease mechanisms, and improve the equitable translation of genomic medicine.

## INTRODUCTION

Large genome-wide association studies (GWAS) and sequencing efforts have identified over 70,000 associations between genetic variants and human diseases and traits^1^. However, despite these advances, a substantial gap between family-based heritability estimates and GWAS-explained variance persists; recent whole-genome sequencing studies suggest that up to 88% of pedigree-based heritability is captured when rare and non-coding variants are included^2^, highlighting the limits of standard SNP arrays for complex trait discovery^3^. This discrepancy, often referred to as the “missing heritability” problem, underscores that a portion of the genetic contribution to complex traits remains unexplained.

Part of this unexplained variance likely reflects ancestry-differentiated variants that remain systematically under-ascertained by discovery cohorts skewed toward a single ancestral background^3^.Most genetic studies have been conducted in individuals of predominantly European-like genetic ancestry, resulting in an underrepresentation of global genetic diversity and limiting the ability to explain disease risk across populations. This limitation has been acutely felt in the use of polygenic risk scores (PRS), whose predictive accuracy decreases as genetic distance from the training population increases, as quantified using principal components or genetic ancestry-inference methods^4,5^. Additionally, population structure, even within relatively homogeneous national biobanks^5,6^, has been shown to impact GWAS results^7,8^. Together, the lack of ancestral diversity and the presence of population structure can reduce statistical power and introduce confounding, thereby hampering the discovery of novel genetic loci using standard GWAS approaches.

Admixture mapping (AM) is a genome-wide association approach that aims to identify susceptibility loci for complex traits by leveraging genetic differences between ancestrally distinct populations represented in reference panels^9^. Thus, AM hinges on local ancestry inference (LAI), which assigns ancestry labels to each chromosomal segment of an individual’s genome. Then, AM tests whether ancestry-associated local ancestry dosages are associated with a phenotype, analogously to GWAS but substituting ancestry dosage for genotype dosage^9^. By exploiting the long-range admixture linkage disequilibrium (LD) of recently admixed populations, ancestry segments tag alleles and haplotypes with population-associated frequencies, allowing loci to be detected with fewer independent tests and greater power^9^. Importantly, the statistical power of AM arises from allele frequency differences across ancestral populations, rather than from differences in disease prevalence itself. When causal variants differ substantially in frequency across ancestry components, local ancestry tracts in admixed individuals can serve as proxies for underlying alleles or haplotypes^10^, thereby generating detectable associations with the phenotype^9^.

Biobank-scale AM has recently become an active area of research, from phenome-wide scans of quantitative traits in the All of Us cohort^11,12^ to conditional fine-mapping frameworks resolving admixture signals to candidate variants across diverse biobanks^13^. These studies have recovered well-established ancestry-differentiated loci, such as the Duffy/*ACKR1* association with white blood cell traits, while uncovering signals not captured by conventional European-centric GWAS, and have shown that conditional analysis can localize a subset of admixture peaks to candidate variants. To the best of our knowledge, however, a comparable biobank-scale AM scan has not been reported in the UK Biobank (UKBB)^14^. Unlike these recent efforts, which were conducted in recently admixed populations, the great majority of non-European UKBB participants are largely single-continental-ancestry migrants rather than products of recent multi-way admixture; the admixture LD is therefore expected to be weaker, and the resolvable haplotype blocks correspondingly longer, than in classically admixed cohorts. Nevertheless, we hypothesized that the scale and phenotypic breadth of the UKBB, combined with eight-way continental ancestry resolution, could provide sufficient power to detect ancestry-associated risk loci for binary disease phenotypes, particularly at loci where causal-allele frequencies differ markedly across ancestral backgrounds.

Here, we performed an AM scan in 415,792 unrelated individuals from the UKBB for 108 phenotypes (Methods, Fig. 1a). We identified 13 ancestry-associated associations, four of which we resolved to a putative causal variant through local-ancestry-conditioned fine-mapping. For these variants, we combined *in silico* variant-effect prediction using Borzoi^15^, a sequence-based deep learning model of gene regulation, with GTEx eQTL annotation, recovering regulatory and coding consequences that were broadly, though not uniformly, concordant across the two approaches. This adds a sequence-level line of functional evidence to admixture mapping signals, complementing the population-level support provided by eQTL data alone. The 13 associations spanned six ancestry components: nine overlapped loci previously reported by GWAS, while four were novel. Notably, the HLA region emerged as a recurrent, ancestry-associated contributor to immune-mediated disease, underlying asthma in South Asian-like ancestry and hypothyroidism in West Asian-like ancestry, and, most strikingly, hypothyroidism risk resolved to distinct background-associated loci: the HLA class II region (6p21.33) in West Asian-like ancestry versus a candidate missense *NKAPL* variant (6p22.2) in African-like ancestry. Together, these results show that local ancestry is not merely a confounder to be corrected for in association studies, but a source of biological information that can reveal how the genetic architecture of a shared clinical phenotype differs across ancestral backgrounds.

**Figure 1.**
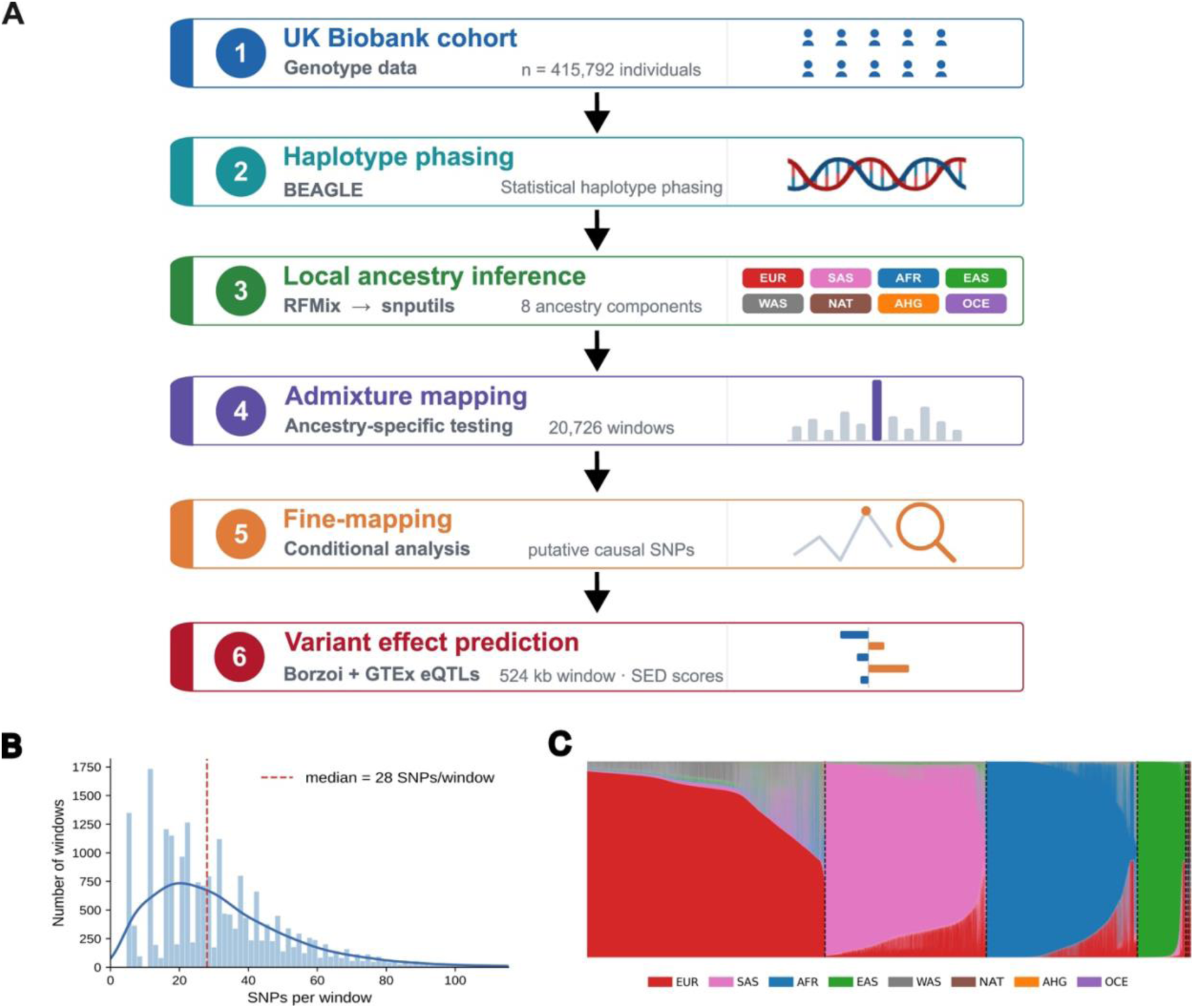
Overview of the multi-ancestry admixture mapping pipeline and dataset characteristics. **A.** Schematic representation of the admixture mapping pipeline. UK Biobank genotype data were phased using BEAGLE, and local ancestry inference (LAI) was performed using RFMix across eight ancestry components (AFR, AHG, EAS, SAS, OCE, EUR, WAS, NAT). LAI outputs were processed using snputils 0.2.1 and used as input for genome-wide admixture mapping analysis. Significant loci were further resolved via fine-mapping analysis, followed by variant effect prediction of the resulting putative causal SNPs. **B.** Density and histogram plot of the number of directly genotyped UK Biobank SNPs per local ancestry window across the genome. **C.** Admixture composition plot for UK Biobank samples with less than 95% estimated European-like ancestry, ordered and color-coded by global ancestry proportion (EUR, SAS, AFR, EAS, WAS, NAT, AHG, OCE).

## METHODS

### UK Biobank data

We considered 108 binary phenotypes extracted from clinical history, cancer registry and family history (Table S1), selected based on their relevance to disease status; non-disease phenotypes were excluded from this analysis. Participants were classified as cases if they received a positive diagnosis at any visit, and as controls otherwise. To assess the independence of the selected phenotypes, we computed pairwise Pearson correlations across all 108 binary phenotypes, finding a mean pairwise correlation of 0.047, indicating that the majority of phenotypes are largely independent of one another (Fig. S1). We used genotype data generated from high-density SNP arrays, including directly genotyped variants (release v2), imputed HLA allelotypes, and imputed genotypes (release v3), all mapped to the hg19 reference^14^. The genotyping platform comprises 658,720 directly genotyped SNPs distributed across the genome, providing high-density marker coverage suitable for fine-scale local ancestry inference and downstream association analyses.

Sample quality control followed UKBB recommendations using their provided sample QC file, `ukb_sqc_v2.txt`. We retained unrelated individuals satisfying the following conditions: 1) inclusion in principal component computation (`used_in_pca_calculation` column); 2) absence of heterozygosity or missingness outliers (`het_missing_outliers` column); 3) no evidence of sex chromosome aneuploidy (`putative_sex_chromo-some_aneuploidy` column). Additionally, we applied a kinship-based exclusion using `plink2 --king-cutoff 0.177` to remove individuals related at first-degree or closer^16^, resulting in a final pool of 415,792 unrelated individuals for downstream analysis.

### Genetic Ancestry Inference

For local ancestry inference, we constructed a reference panel using samples from the 1,000 Genomes Project^17^, HGDP^18^, and SGDP^19^. We inferred unsupervised ancestry clusters using ADMIXTURE^20^ at K=8 and retained only individuals with ≥95% assignment probability to a single cluster. Clusters were labeled based on ancestral origin^21^ as: African-like ancestry tracts (AFR), East Asian-like ancestry tracts (EAS), South Asian-like ancestry tracts (SAS), Oceanian-like ancestry tracts (Australo-Papuan) (OCE), European-like ancestry tracts (EUR), West Asian-like ancestry tracts (WAS), African Hunter Gatherer-like ancestry tracts (AHG), and Indigenous American-like ancestry tracts (NAT). We used worldwide references because the biobanks contain individuals admixed from populations around the world, so no particular admixture model can be assumed a priori for a given subpopulation. Commonly used simplified models, for example, a three-way NAT-EUR-AFR model for Latin American populations, have been shown to be misspecified, given documented EAS/OCE and SAS admixture in some of these groups^22,23^. The final reference panel (Table S2) comprised 1,380 individuals across eight clusters and 605,169 SNPs. All samples were phased jointly using *beagle*^24^ with default parameters. Local ancestry inference was performed using RFMix, a discriminative conditional random field approach parameterized with random forests^25^, by executing the command `rfmix -f $query -r $ref -m $sample_map -g $genetic_map -o $output --n-threads=8 --chromosome=${chr}`. Summary statistics are reported in Table S3. RFMix incorporates the supplied genetic recombination map so that local variation in recombination rate inform ancestry-state transitions^25^.To assess inference stability in regions of complex LD, we performed a targeted quality-control analysis of the HLA locus by quantifying ancestry assignment frequencies across the HLA locus and comparing them with those across the rest of the genome, confirming the absence of systematic ancestry overrepresentation (Table S4). To assess the robustness of our local ancestry calls to the choice of inference method, we compared RFMix and Gnomix^21^ on chr6 and chr22 in samples with low EUR-like ancestry (n=21,058), observing high concordance of per-window ancestry assignments (Cohen’s κ = 0.90 on both). Concordance was slightly lower within the HLA region (Cohen’s κ = 0.81) yet remained high overall.

### Admixture mapping association analysis

Among common AM frameworks, we adopted the ancestry-aware approach, which tests each ancestry-component dosage separately, as it can reveal effects masked in joint testing^9^. We first converted the LAI output in MSP format to variant calling format (VCF) using snputils (0.2.1)^26^. Then, we performed AM by fitting an ancestry-associated logistic regression for each phenotype, ancestry, and local ancestry window using plink2 (`--glm` option)^16^

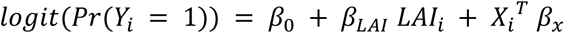

where *Y_i_* is the binary phenotype of individual i, *LAI_i_* ∈ {0, 1, 2} is the ancestry-associated local ancestry dosage at the tested window (number of haplotype copies assigned to the local ancestry, as inferred by RFMix), *X_i_* is the vector of covariates (i.e. age, sex, BMI, and global ancestry proportions), *β*_0_ is the intercept, *β_LAI_* is the local-ancestry effect of interest, and *β_x_* is the vector of covariate effects. Global ancestry proportions were preferred over principal components as covariates, as PCA in admixed populations can produce higher-order components that capture local genomic features rather than genome-wide ancestry, potentially inducing collider bias and spurious associations^27^. The European global ancestry proportion was omitted from *X_i_* to avoid perfect collinearity, because global proportions sum to one. All continuous covariates were standardized to zero mean and unit variance. When standard maximum-likelihood logistic regression failed to converge, Firth-penalized logistic regression was applied.

Association testing was performed independently for each *ancestry - phenotype* combination, yielding *m* tests per combination, where *m* is the number of genome-wide local-ancestry windows. To control for genomic inflation, we computed the genomic inflation factor λ for each ancestry–phenotype combination and retained only analyses with 0.9 ≤ λ ≤ 1.1^28^ (QQ plots, Fig. S2–14). Within each retained combination we controlled the false discovery rate across the *m* genome-wide windows using the Benjamini-Yekutieli (BY) procedure^29^, which is valid under arbitrary dependence. We required that the number of declared windows k satisfy 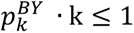, thereby restricting the expected number of false discoveries to at most one per combination. Finally, because local ancestry dosages are near-identical between neighboring windows, genuine tract-driven associations are expected to span multiple windows; we therefore declared a signal genome-wide significant only where ≥5 spatially contiguous windows jointly met these criteria, a requirement far shorter than the expected tract lengths in this cohort, and one that can only remove signals, never add them.

### Fine-mapping analysis conditioning on local ancestry

Local ancestry tracts identified by admixture mapping often span multiple megabases and may harbor multiple candidate variants in LD with the causal variants. To refine these signals and identify variants that may drive the ancestry-dependent associations, we implemented a conditional analysis-based fine-mapping procedure.

For each admixture mapping hit, we performed a genome-wide association analysis restricted to the genomic region spanned by the hit, fitting an ancestry-associated logistic regression model that conditions on local ancestry:

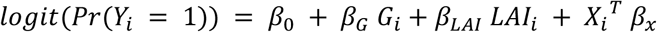

Where *G_i_* ∈ {0, 1, 2} is the genotype dosage at the tested SNP for individual i, *LAI_i_* is the local ancestry dosage in the region, *X_i_* is the vector of covariates (i.e., age, sex, BMI, global ancestry proportions), *β_G_* is the SNP effect of interest, and *β_LAI_* captures any ancestry effect independent of the tested SNP. Association testing was performed using plink2 (`--glm` option with firth fallback). Under this approach, if an LAI segment serves as a proxy for an unmeasured causal variant, then controlling for that causal variant should substantially attenuate the local ancestry signal.

We selected candidate SNPs as putatively causal based on three simultaneous criteria evaluated in the ancestry-associated conditional model (Section 2.4.1). The SNP had to reach locus-wide significance with a BY corrected p-value below 0.05 and have an odds ratio consistent with the direction of the AM signal. Concurrently, the local ancestry signal had to become non-significant when conditioned on the SNP, with the BY-FDR-corrected LAI p-value exceeding 0.05. This criterion ensures that the selected SNPs contain sufficient information to explain the LAI signal, suggesting that they are likely tagging the causal variants driving the AM association. Among the extracted candidates, we selected the variant with the largest odds ratio as a representative tag for the signal; we interpret it as a candidate rather than the definitive causal variant.

### Functional annotation of fine-mapped variants

Fine-mapped variants were annotated using Ensembl^30^ for genomic context, the GWAS Catalog^31^ to check for prior association, Borzoi^15^ to predict variant effects on gene regulation, and GTEx v10^32^ for annotated eQTL. Borzoi takes as input a ∼524 kb genomic sequence window centered on each variant and predicts a set of functional genomic tracks (e.g., RNA-seq, ATAC-seq, ChIP-seq); variant effects are quantified as SED (SNP Expression Difference) scores, defined as the predicted difference in expression of a specific target gene between the alternative and reference allele sequences. Protein domain architecture, disordered regions, and post-translational modification sites were retrieved from UniProt^33^. For the frequency extraction, we used GnomAD^34^.

## RESULTS

### Admixture mapping analysis identifies ancestry-associated phenotype associations in UKBB individuals

We first performed LAI in 415,792 individuals from UKBB, across 20,726 genomic windows using *RFMix*^25^ (Methods). These windows covered the autosomes, with a median of 28 SNPs per window (Fig. 1B). Then, global ancestry proportions were estimated as the fraction of genome-wide local ancestry windows assigned to each ancestry component.

As expected, we found that >95% of individuals were at least 50% of EUR ancestry (Fig. 1C). Nevertheless, we found that 8,965 individuals were predominantly SAS; 8,422 were predominantly AFR; 2,742 were predominantly EAS; 151 were predominantly NAT; and 122 were predominantly WAS, where an individual was considered predominantly of one ancestry if their global ancestry proportion exceeded 50%.

Next, we selected 108 binary phenotypes (Methods; Table S1) and tested the association between local ancestry and each phenotype across genomic windows for every ancestry–phenotype combination (Methods). This analysis identified 13 significant ancestry–phenotype associations, spanning 333 genomic windows (8,157 SNPs) and mapping to nine distinct genomic loci, as several phenotypes reflect overlapping clinical codings (Table S5). To evaluate the statistical validity of these results, we produced quantile-quantile (QQ) plots and quantified genomic inflation (Fig. S2-14). This analysis showed controlled inflation, with coefficients ranging from 0.95 to 1.10 in these cases (Table S5), suggesting that our statistical model can reliably produce p-values.

A total of nine regions contained genes previously associated with the tested phenotype by GWAS strategies, and four were considered new findings. Among regions overlapping with previous findings, we identified associations with AFR-like ancestry spanning four phenotypes, hypothyroidism, other hypothyroidism, myocardial infarction and heart attack, across 51 regions; with SAS-like ancestry spanning two asthma and asthma-related phenotypes across 123 regions; and with WAS-like ancestry spanning three phenotypes, gastroesophageal reflux, hypothyroidism and hypothyroidism-related conditions - across 132 regions. Among novel associations, we identified dermatitis and atrial fibrillation/flutter with EAS-like ancestry (13 regions), atrial fibrillation/flutter with NAT-like ancestry (8 regions), and angina pectoris with EUR-like ancestry (6 regions).

We subsequently narrowed down the regions via fine-mapping analysis (Methods), resulting in a putative causal SNP for four associations (Fig. 2B, Table S6, Fig. S15-20), for which we performed variant effect prediction (VEP) analysis (Methods).

**Figure 2.**
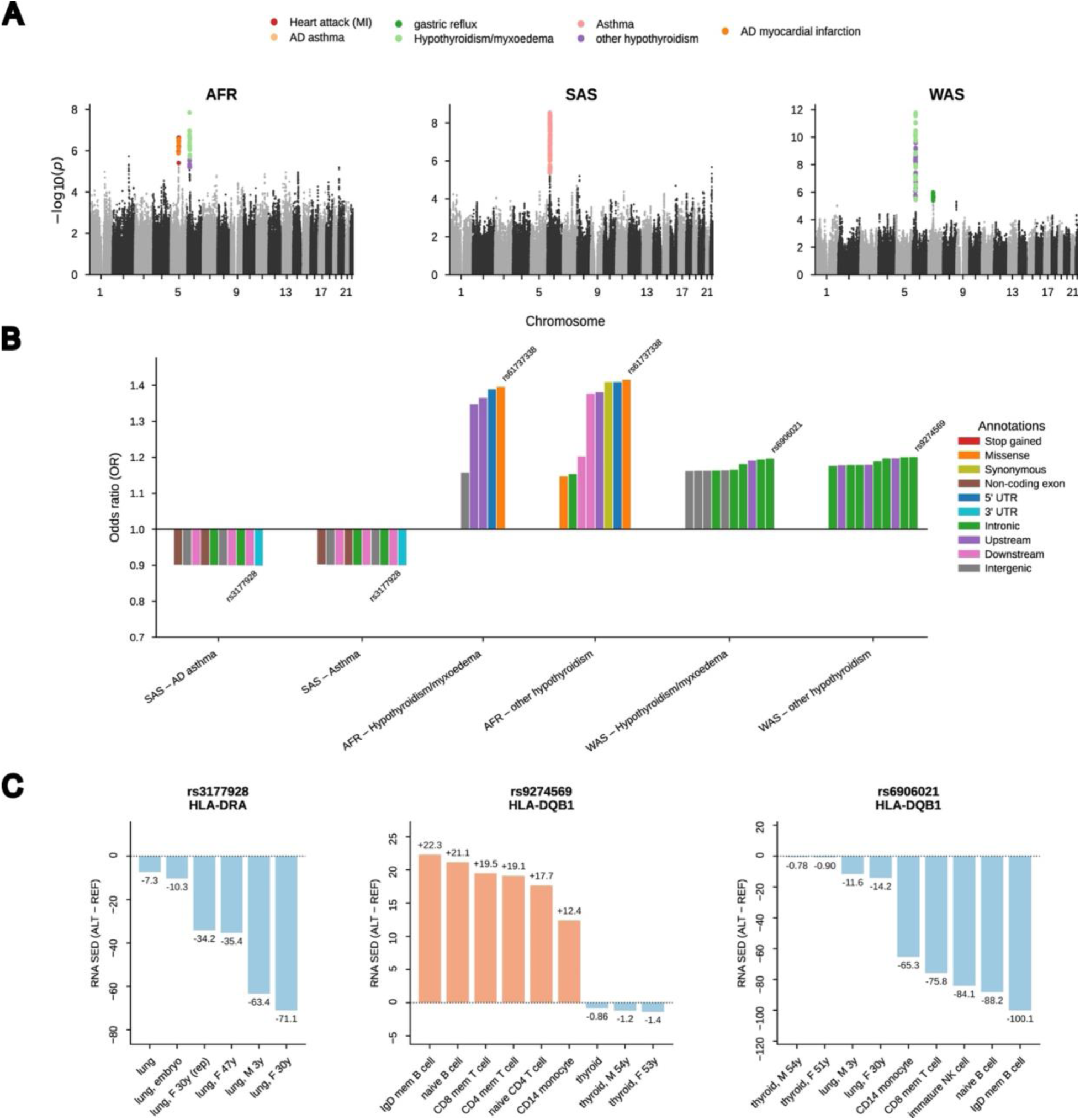
Fine-mapping results and genome-wide association signals for replicated ancestry-associated loci. **A.** Manhattan plots for the three ancestries yielding replicated admixture mapping signals: AFR-like, SAS-like, and WAS-like. The x-axis shows chromosomal position, and the y-axis shows −log₁₀(p-value). Colored points indicate genome-wide significant hits, color-coded by phenotype as shown in the legend. **B.** Functional annotation of the four putative causal SNPs identified by conditional analysis across all significant ancestry-phenotype combinations. The bar chart shows the distribution of the 10 most impactful variant annotations stratified by ancestry-phenotype association, with the putative causal SNPs selected. **C.** Borzoi-predicted regulatory effects (SED score) for the four fine-mapped SNPs with a coherent functional signal, shown as the top affected tracks per variant, colored by direction of effect (decrease in blue, increase in orange). rs61737338 is omitted, as no coherent signal was found.

### Ancestry-associated risk loci across phenotypes with inconclusive fine-mapping evidence

Three phenotype-ancestry associations did not yield any putative causal variant and therefore could not be resolved to a specific locus: one gastro-esophageal reflux signal in WAS-like ancestry, and two overlapping cardiovascular signals (heart attack and myocardial infarction) in AFR-like ancestry. The former consisted of a 1.9 Mb segment on chromosome 7p12.1 showing a higher risk of gastro-esophageal reflux in WAS-like ancestry (OR = 1.16, 95% CI: 1.09–1.23). Across this interval, linkage disequilibrium (LD) is dispersed into small blocks rather than concentrated in one, and no SNP reached significance after BY correction (Fig. S21), likely reflecting a shortage of individuals carrying the local ancestry tract at this specific locus rather than an unresolvable signal. This region includes genes such as *LINC01446*, *VSTM2A*, and *POM121L12*. Notably, *LINC01446* has been previously implicated as an oncogenic poor-prognosis marker in gastric cancer in Chinese populations^35^, suggesting a potential functional link between this locus and upper gastrointestinal pathology. In addition to this association, we also identified a protective association for heart attack in a 2.3 Mb segment on chromosome 5q14.3 for AFR-like ancestry (OR = 0.53, 95% CI: 0.42–0.68). As with the WAS locus, LD here is discontinuous, with no SNP surviving BY correction (Fig. S22). This region contains *MEF2C* and *TMEM161B-DT*. *MEF2C* is a cardiac transcription factor; it has been linked to congenital double outlet right ventricle in Chinese populations^36^, although that structural developmental phenotype differs from the adult ischemic event captured here. In addition, another concordant protective association was observed for myocardial infarction in the same region, showing the same discontinuous LD and null conditional signal (Fig. S23); because heart attack and myocardial infarction reflect largely overlapping phenotype definitions in the UKBB, this recovery is expected and should be read as internal consistency rather than independent replication.Taken together, these loci represent ancestry-associated associations for which conditional fine-mapping did not isolate a single putative causal variant, most plausibly reflecting a shortage of individuals carrying the relevant local ancestry tract at each locus.

### Fine-mapping analysis in the HLA region identifies candidate regulatory variants for asthma

Among our admixture mapping analysis hits we identified two overlapping protective associations for asthma phenotypes (Asthma and AD Asthma) in the HLA class I and II region (6p21.33–6p22.1) for SAS-like ancestry (OR = 0.91–0.92, 95% CI: 0.88–0.94), consistent with prior GWAS reports ^37,38^ (Fig. 2A, Table S5) Fine-mapping analysis of the corresponding genomic regions led us to SNP rs3177928 (Fig. 2B).). Across all two loci, the single most significant SNP after BY correction is not among the fine-mapped candidates (Figs. S24-S25), consistent with our conditional criterion (Methods) selecting the variant that explains the local-ancestry signal rather than simply the most significant one. Population-level allele frequencies (Table S6) or rs3177928, shows that the highest frequency is observed in non-admixed European populations (14.84%), with the South Asian frequency closely following (14.05%).

Inspection of the genomic annotation of these loci revealed that rs3177928is found in the 3’ UTR region of *HLA-DRA*. The annotation of that variant suggests it could be involved in post-transcriptional regulation of *HLA-DRA* expression, potentially modulating antigen presentation and immune responses relevant to asthma susceptibility. Moreover, this gene has previously been associated with asthma in Japanese populations^39^. Notably, the same genomic window produced a significant association for EUR-like ancestry (OR = 0.94, 95% CI: 0.93–0.96), which was excluded due to a large genomic inflation factor of *λ* = 1.30 in that case (Methods).

Next, we used complementary variant effect prediction methods to explore the functional implications of this putative causal variant. Borzoi *in silico* prediction for rs3177928 showed a consistent decrease in predicted *HLA-DRA* expression across multiple independent lung tracks (SED = −71.1 to −7.3, across several adult and pediatric samples, Fig. 2C). Borzoi and GTEx implicate different target genes at this locus: Borzoi predicts decreased *HLA-DRA* expression, whereas the GTEx lung eQTLs act on other genes, namely a decrease in *HLA-DQB2* expression (NES = −0.55, P = 4.6 · 10⁻¹⁵) and in *HLA-DRB9* expression (NES = −0.52, P = 4.0 · 10⁻¹⁴).

### WAS- and AFR-associated hypothyroidism associations refined to HLA region, and NKAPL

Both hypothyroidism and other hypothyroidism showed overlapping signals, partially covering the *HLA* class I and II regions (6p21.33), with ORs of 1.10 (95% CI: 1.07–1.14) and 1.12 (95% CI: 1.08–1.15), respectively, for WAS-like ancestry. The class I region harbors *HCP5*, a gene previously implicated in autoimmune thyroid disorders in European populations^40^. More specifically, an intronic variant in *HCP5* has been associated with both susceptibility to and age of onset of Graves’ disease in UK and Polish cohorts^41^, while other variants have been linked to thyroid peroxidase antibody levels and clinical thyroid disease susceptibility^40^. It is worth noting that this prior *HCP5* evidence derives from European cohorts and largely concerns autoimmune thyroid disease including hyperthyroid Graves’ disease, distinct from the hypothyroidism captured here; moreover, our fine-mapping did not implicate *HCP5* but resolved instead to the neighboring class II region. Fine-mapping analysis of the region led us to SNPs rs9274569 and rs6906021, both found in the class II region (Fig. 2B). As at the SAS asthma locus, LD across this interval is dispersed into small, discontinuous blocks, with rs9274569 and rs6906021 recovered as the fine-mapped, concordant candidates against this background (Figs. S26–S27). The former is an intronic SNP found in *HLA-DQB1*, while the latter maps to an intergenic region *between HLA-DQA1* and *HLA-DQB1*. Consistent with our results, *HLA-DQB1* has been previously associated with hypothyroidism^42,43^, with *HLA-DQA1* specifically implicated in Taiwanese populations^42^. Population-level allele frequencies (Table S6) are consistent with the WAS-associated signal: rs9274569 reaches its highest frequency in WAS populations (54.76%), as does rs6906021 (65.75%).

Borzoi RNA prediction for the identified variant in rs9274569 showed an upregulation in *HLA-DQB1* expression across lymphoid and myeloid RNA tracks (SED = +12.4 to +22.3, Fig. 2C); this is opposite to the GTEx Whole Blood eQTL for *HLA-DQB1* (NES = −0.57, P = 2.3·10⁻⁷⁴), but matches the direction of its paralog *HLA-DQB2* in whole blood tissue (NES = +0.70, P = 1.3 · 10⁻⁷⁰). On the other hand, Borzoi RNA predictions for thyroid-specific tracks showed a small decrease in *HLA-DQB1* expression (SED = −1.4 to −0.86, Fig. 2C), matching GTEx Thyroid eQTL for *HLA-DQB1* itself (NES = −0.68, P = 2.1 · 10⁻⁶⁰), but opposite to its paralog *HLA-DQB2* in the same tissue (NES = +0.74, P = 1.7 · 10⁻⁶⁹).

The identified variant in rs6906021 showed a consistent decrease in *HLA-DQB1* expression across lymphoid/myeloid tracks (SED = −100.1 to −65.3, Fig. 2C) and, more modestly, in lung (SED = −14.2 to −11.6, Fig. 2C) and thyroid (SED = −0.90 to −0.78, Fig. 2C). These results were consistent with the GTEx eQTL for *HLA-DQB1* in whole blood tissue (NES = −0.55, P = 5.5 · 10⁻⁷⁷). GTEx eQTLs, however, also point to upregulation of its paralog *HLA-DQB2*, which is increased in thyroid (NES = +0.73, P = 1.4 · 10⁻⁷³), whole blood (NES = +0.64) and muscle-skeletal tissue (NES = +0.83).

Beyond the HLA class II signal identified in WAS-like ancestry, our analyses revealed a distinct genomic locus associated with hypothyroidism risk in AFR-like ancestry, highlighting ancestry-associated genetic architectures underlying the same phenotype. We identified risk associations with both hypothyroidism/myxoedema (OR = 1.14, 95% CI: 1.09–1.19) and other hypothyroidism (OR = 1.12, 95% CI: 1.07–1.17) in overlapping ∼2 Mb segments on chromosome 6p22.2. Thus, whereas the WAS-like association localized to the HLA class II region at 6p21.33, the AFR-like signal mapped to a neighboring but separate region on chromosome 6p22.2, outside the classical HLA class II genes.

Fine-mapping analysis of the region led us to rs61737338, a missense variant in *NKAPL* (p.Ser23Phe; SIFT deleterious) that abolishes an annotated phosphoserine within a disordered, low-complexity N-terminal region, potentially affecting post-translational regulation of the protein rather than its fold (Fig. 2a). This should be interpreted cautiously: Borzoi predictions and GTEx eQTLs were not concordant for this variant, and its relatively high population frequency is compatible with a tolerated polymorphism, so we regard it as a candidate rather than an established driver. Consistent with this caution, the BY-corrected signal at this locus is markedly weaker than at the WAS-like HLA class II locus (peak −log10 BY p ≈ 15 vs. ≈150; Figs. S28, S29 vs. S26, S27), against a denser, more block-structured LD background than seen at the other resolved loci. However, this locus lies immediately adjacent to the MHC, a region robustly associated with autoimmune hypothyroidism^44^. Population-level allele frequencies (Table S6) are consistent with the AFR-associated signal, with rs61737338 reaching its highest frequency in African populations (6.8%).

### New ancestry-associated associations without resolved causal variants

Among the 13 associations, we identified four new ancestry-associated risk associations (Fig. 3A). These were considered novel after querying the GWAS Catalog and the literature for prior associations at each locus–phenotype pair (Methods). For NAT-like ancestry tracts, we found a risk association with atrial fibrillation and flutter in a 917 kb segment on chromosome 6p22.3 (OR = 4.69, 95% CI: 2.48–8.90). The segment spans *DCDC2*, *NRSN1*, *ISG20L2P1*, not established atrial-fibrillation loci. For EAS-like ancestry, a 4.3 Mb region on chromosome 20p11.1 showed risk association with atrial fibrillation (OR = 1.49, 95% CI: 1.28–1.74). This interval contains *HM13*, *BCL2L1*, *TPX2*, *TTLL9*, none previously linked to atrial fibrillation. Among our EUR-like ancestry analyses, we identified a 504 kb segment on chromosome 10p14 associated with angina pectoris (OR = 1.33, 95% CI: 1.18–1.49), containing genes *ITIH5* and *TAF3*. A single-window signal for angina pectoris on chromosome 6p22.3 (OR = 1.32, 95% CI: 1.17–1.48) did not meet our ≥5-contiguous-window criterion (Methods; Table S5).

**Figure 3.**
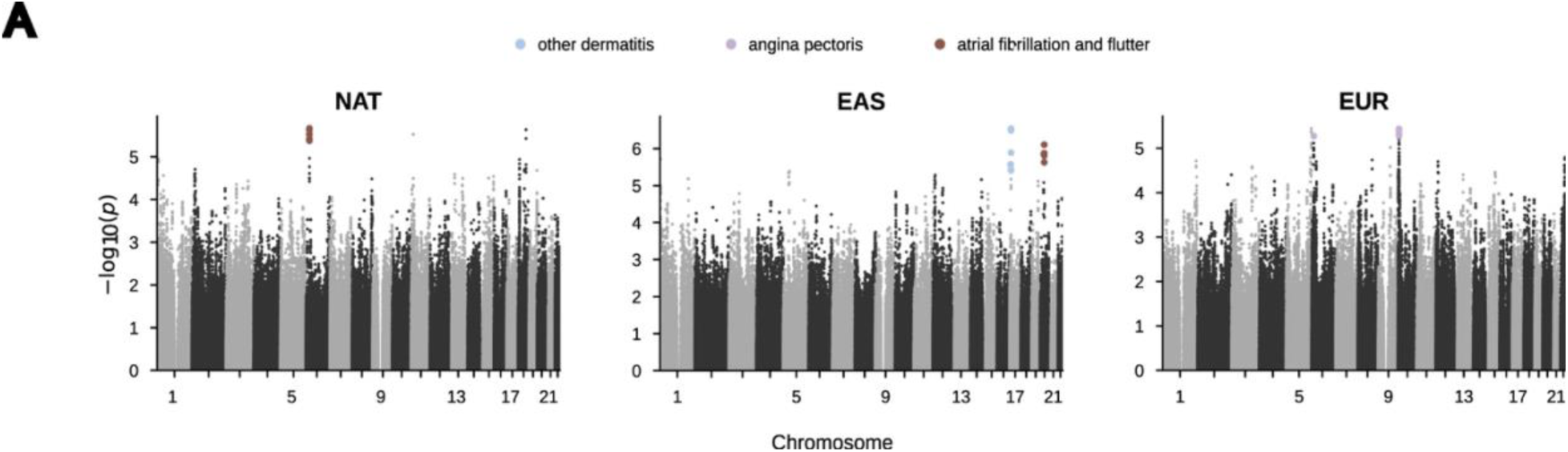
Genome-wide association signals for novel ancestry-associated loci. **A.** Manhattan plots for the three ancestries yielding novel admixture mapping signals: NAT-like, EUR-like, and EAS-like. The x-axis shows chromosomal position, and the y-axis shows −log₁₀(p-value). Colored points indicate genome-wide significant hits, color-coded by phenotype as shown in the legend.

Although these four associations represent novel ancestry-associated signals not previously reported in the literature, fine-mapping did not resolve a putative causal variant in any of these regions, leaving the underlying mechanisms unresolved: LD across all four loci is dispersed into small, discontinuous blocks and no SNP reached significance after BY correction (Figs. S30–S33), a pattern most consistent with a shortage of local-ancestry tract carriers at each locus rather than an unresolvable biological signal. The magnitude of the NAT-like atrial fibrillation signal is particularly striking but rests on a wide confidence interval, which we owe to the smaller sample size for that ancestry. Finally, among our EAS-like ancestry analysis, we identified a 759 kb risk locus on chromosome 17p13.1 associated with dermatitis (OR = 1.41, 95% CI: 1.24–1.61). This region contains *NTN1*, previously associated with malignant melanoma^45^, but this is the first time an association with dermatitis is reported to the best of our knowledge.

## DISCUSSION

Despite the predominantly European composition of the UKBB, our framework combining both admixture and fine-mapping analysis, which directly models the ancestral origin of genetic variants, identified 13 admixture mapping associations across 333 genomic windows spanning six ancestry components, pinpointing in four genomic windows their corresponding putative causal variants. Nine of these hits were previously reported by conventional strategies^35–44^ supporting the validity of local-ancestry-based methods while also identifying the ancestral background driving each association; the remaining four appear novel: atrial fibrillation in NAT-and EAS-like ancestries, dermatitis in EAS-like ancestry, and angina pectoris in EUR-like ancestry.

Local-ancestry-conditioned fine-mapping resolved four associations to a putative causal variant; complementary variant-effect prediction then added a functional layer that was broadly, though not uniformly, concordant across the two approaches. Combining local ancestry information with variant-level analyses helped narrow loci that single-population studies might overlook. Conditioning on local ancestry dosage during fine-mapping was central to this step. We emphasize that our procedure identifies the variant that best statistically accounts for the local ancestry signal under our criteria, not necessarily the causal variant itself; in regions of strong admixture and background LD the true causal allele may remain unmeasured or may reflect a haplotype-level combination of variants rather than a single site^10^, so further validation in sequencing-based case/control studies remains essential.

Fine-mapping of the SAS-like asthma–HLA signal identified rs3177928, a 3’ UTR variant in *HLA-DRA*, as the putative causal variant, consistent with the established role of this gene in antigen presentation and asthma susceptibility^37–39^. Borzoi predicts decreased *HLA-DRA* expression in lung tissue at this variant, while GTEx lung eQTLs implicate distinct downstream targets *(HLA-DQB2* and *HLA-DRB9*), suggesting that the regulatory mechanism at this locus may act through multiple effector genes. The lower asthma prevalence reported in Asian-than European-ancestry US populations^46^ is consistent with the hypothesis that HLA-region risk is modulated by population-associated factors beyond allele frequency alone. The WAS-like hypothyroidism signal localized to the *HLA* class II region and fine-mapped to two candidate variants at *HLA-DQB1* (rs9274569, rs6906021), a gene with prior support in hypothyroidism^43^. For both, predicted regulatory effects were larger in immune than in thyroid tracks and consistently opposite to the paralog *HLA-DQB2* (Results), pointing to an immune-rather than thyroid-resident mechanism, though paralog-level assignments at HLA should be read cautiously given known mapping and imputation difficulties. By contrast, the AFR-like hypothyroidism signal mapped to a separate neighboring locus (6p22.2) and to a missense *NKAPL* variant (rs61737338) for which Borzoi and GTEx were discordant and whose high frequency is compatible with a tolerated polymorphism; we regard it as a candidate only. Hypothyroidism thus exemplifies ancestry-associated heterogeneity, the same phenotype arising through distinct loci in different backgrounds, and, with the asthma findings, marks the *HLA* region as a recurring, ancestry-dependent contributor to immune-mediated disease. The four novel associations noted above remained unresolved by fine-mapping; despite this, they illustrate the potential of ancestry-aware analyses in underrepresented populations and will require replication and functional follow-up.

Several limitations should be noted. The UKBB is predominantly European: the reference panels and the counts of predominantly WAS-like (122) and NAT-like (151) participants are far smaller for WAS-, AHG-, and OCE-like ancestries, limiting power. Because most non-European participants are largely unadmixed, admixture LD is weaker and longer-range than in classically admixed cohorts, most plausibly why 9 of the 13 associations went unresolved, and a caveat for effect sizes resting on small tract-carrier counts. Resolution is further constrained by LAI accuracy, imputation quality, and complex LD, notably at HLA, where ancestry proportions diverge little from the genome-wide background (Table S4), yet reduced LAI accuracy may still bias results. Within each fine-mapped region we also examined SNPs with lower p-values than the selected candidate. These SNPs were tested in the same ancestry-associated model used for fine-mapping (Methods), which already conditions on the LAI dosage at the locus; however, when we in turn conditioned on each of these SNPs, the LAI term remained significant, showing that, unlike the selected candidate, they do not explain the local-ancestry admixture signal. These non-explanatory, SNPs were themselves largely more frequent in European (non-Finnish; 10.5–58.6%) and Amish (12.2–59.2%) reference panels than in the ancestry carrying the signal (2.96–50.9%), consistent with them tagging EUR-heavy background LD (Fig. S21-33) rather than the admixture signal itself; the sole exception falls precisely in the region where the EUR-like association itself had been excluded as inflated yet flagged as plausible. Finally, our functional interpretation rested on computational predictions and public datasets, and ancestry-diverse replication cohorts remain scarce; both should improve as these resources grow.

In summary, admixture mapping coupled with ancestry-informed fine-mapping uncovered ancestry-associated disease loci across a broad phenotypic range, with the contrasting AFR-and WAS-like hypothyroidism signals illustrating how one clinical phenotype can arise through distinct loci in different backgrounds. Future work should prioritize replication in ancestry-matched biobanks, functional characterization of the candidate variants, and integration with transcriptomic and epigenomic data. These findings reinforce that modelling local ancestry is a source of biological information, not merely a confounder, for disease-gene discovery and the equitable translation of genomic medicine.

## DATA AVAILABILITY

The UKBB raw data can be obtained through a data access application available at https://www.ukbiobank.ac.uk. The 1000 Genomes reference panel is available at ftp://ftp.1000genomes.ebi.ac.uk/vol1/ftp/release/20130502/. The Human Genome Diversity Project dataset is available in a GitHub repository at: https://www.internationalgenome.org/data-portal/data-collection/hgdp.

## CODE AVAILABILITY

The code employed in this research is available in a GitHub Repository at: https://github.com/Danariki99/Admixture_mapping.

## ACKNOWLEDGEMENTS

This work uses data provided by patients and collected by the NHS as part of their care and support. We thank the UK Biobank and its participants and the NHS. This research was conducted using the UK Biobank Resource under Application Number 24983.

## AUTHOR CONTRIBUTIONS

AGI, DMM, and SMG conceived the study; GV, AGI, and MAR conducted the local ancestry inference; RS conducted the admixture mapping, fine-mapping, and variant effect prediction analyses, with assistance from SMG, DB and CF; RS and JA wrote the manuscript, with input from SMG; AGI, JA, DMM, MAR, SMG, AS, DB, and SDC.

## ADDITIONAL INFORMATION

**Supplementary information** All supplementary tables can be found in the Supplementary_tables.xlsx file.

