## Supplementary Figures for "Multi-ancestry admixture mapping reveals ancestry-associated disease loci in the UK Biobank"

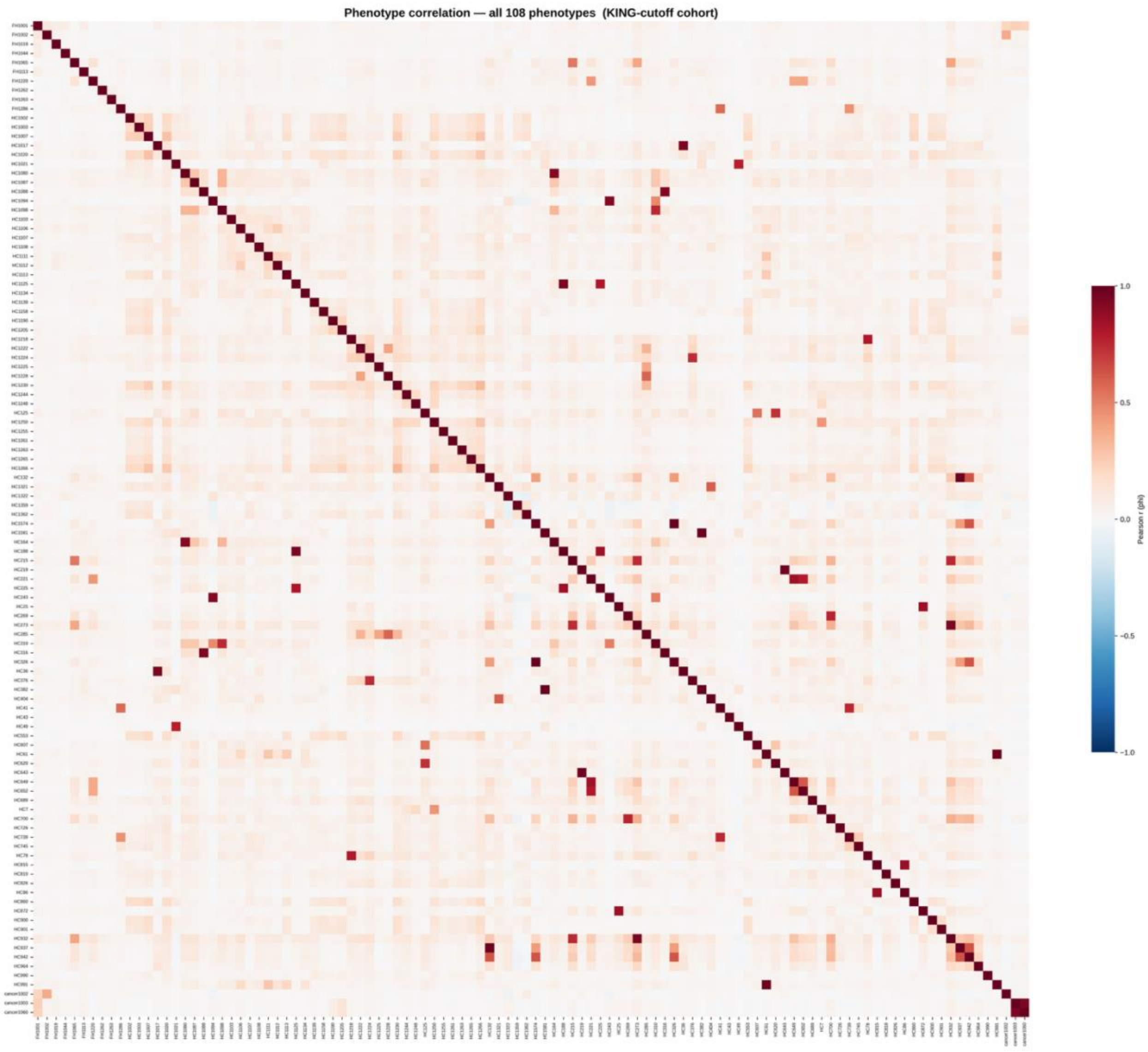


**Supplementary Figure 1.** Pairwise Pearson correlation matrix across all binary phenotypes. Heatmap of pairwise Pearson correlation coefficients computed across all 108 binary phenotypes analyzed in this study (mean pairwise r = 0.047), indicating that the majority of phenotypes are largely independent of one another.

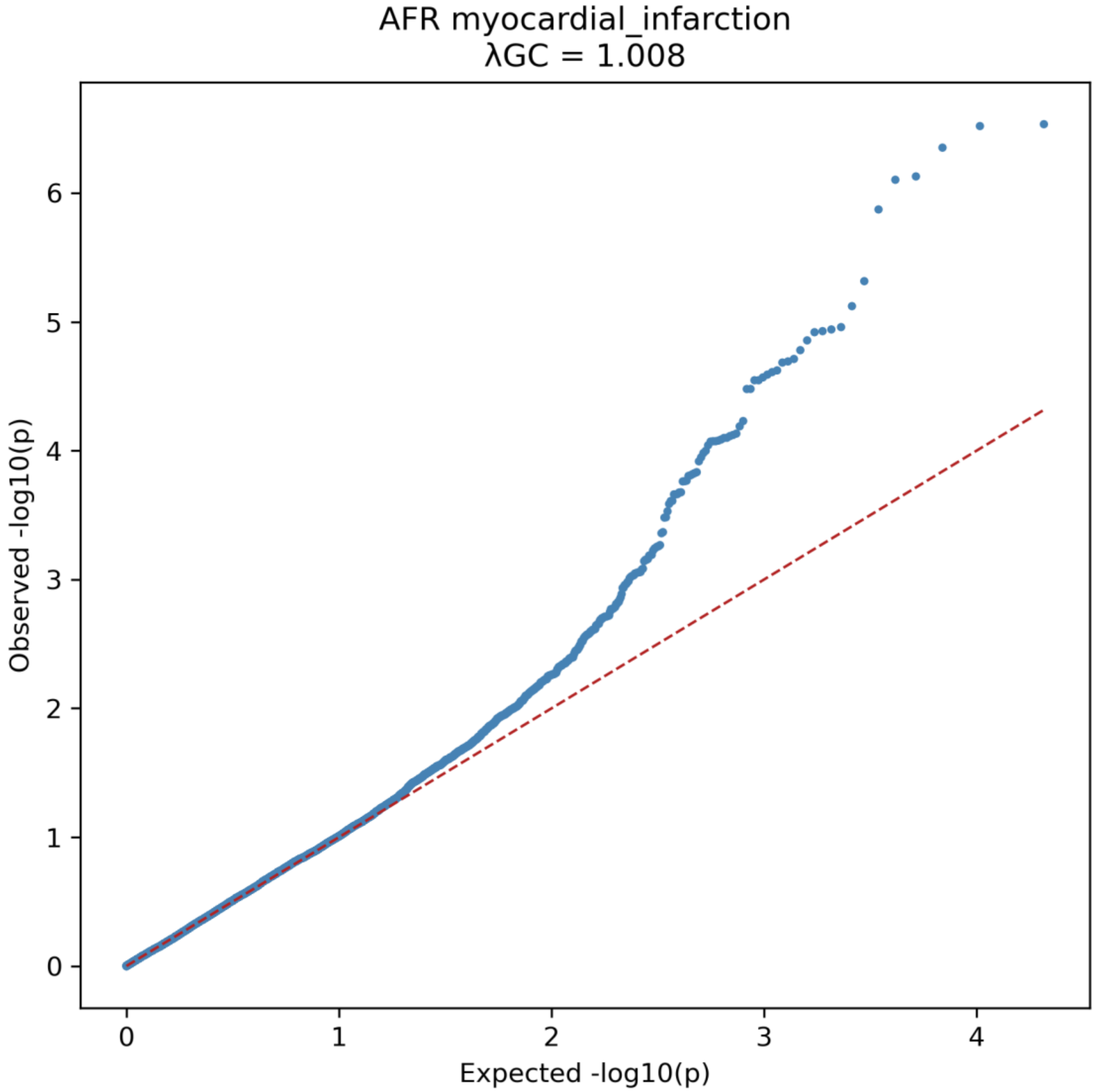


**Supplementary Figure 2**. Quantile–quantile plot for AD myocardial infarction in the AFR ancestry group. Observed versus expected −log10(p) values are shown, with λGC = 1.008. The red dashed line indicates the null expectation under no association.

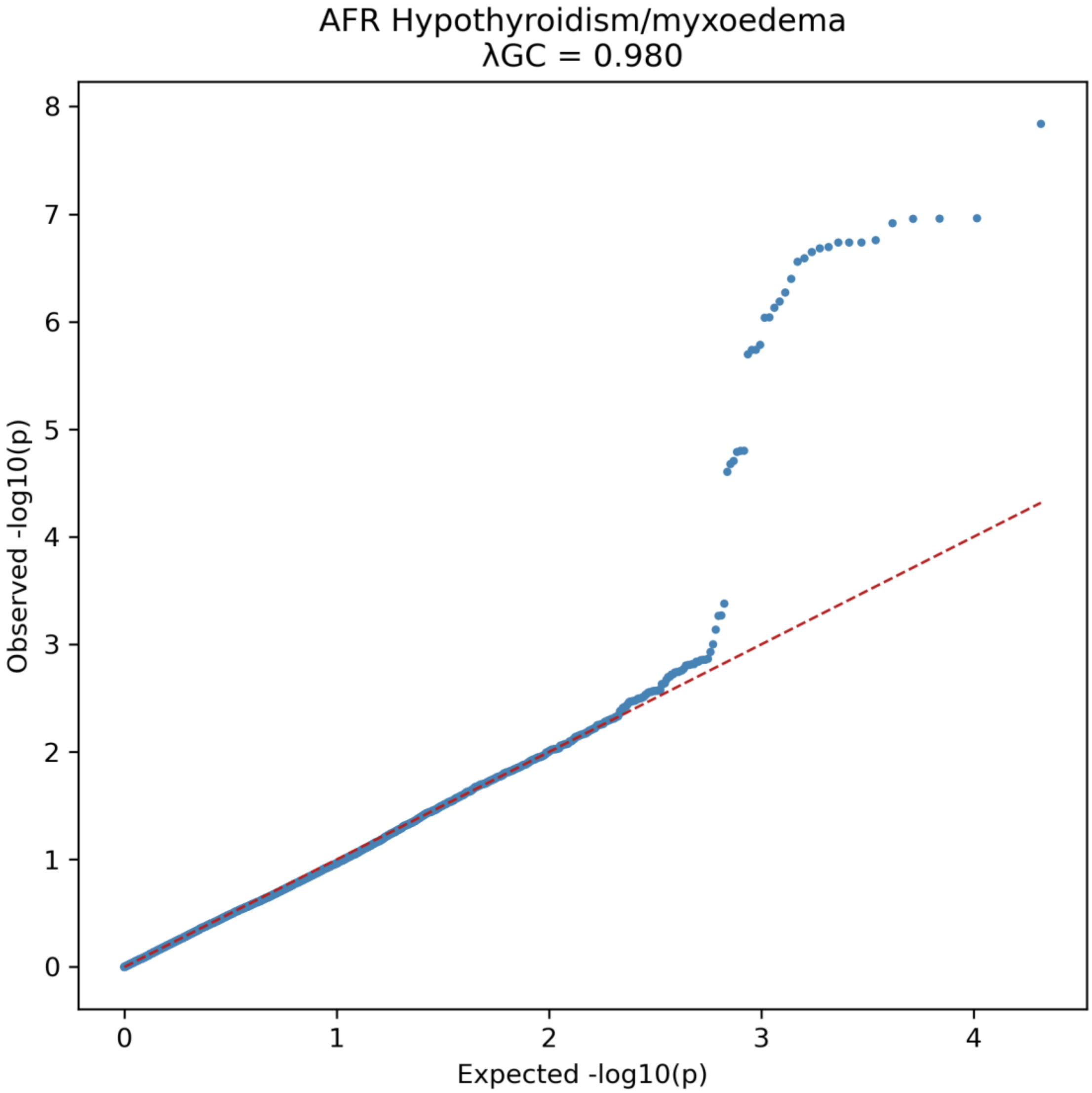


**Supplementary Figure 3**. Quantile–quantile plot for Hypothyroidism/myxoedema in the AFR ancestry group. Observed versus expected −log10(p) values are shown, with λGC = 0.980. The red dashed line indicates the null expectation under no association.

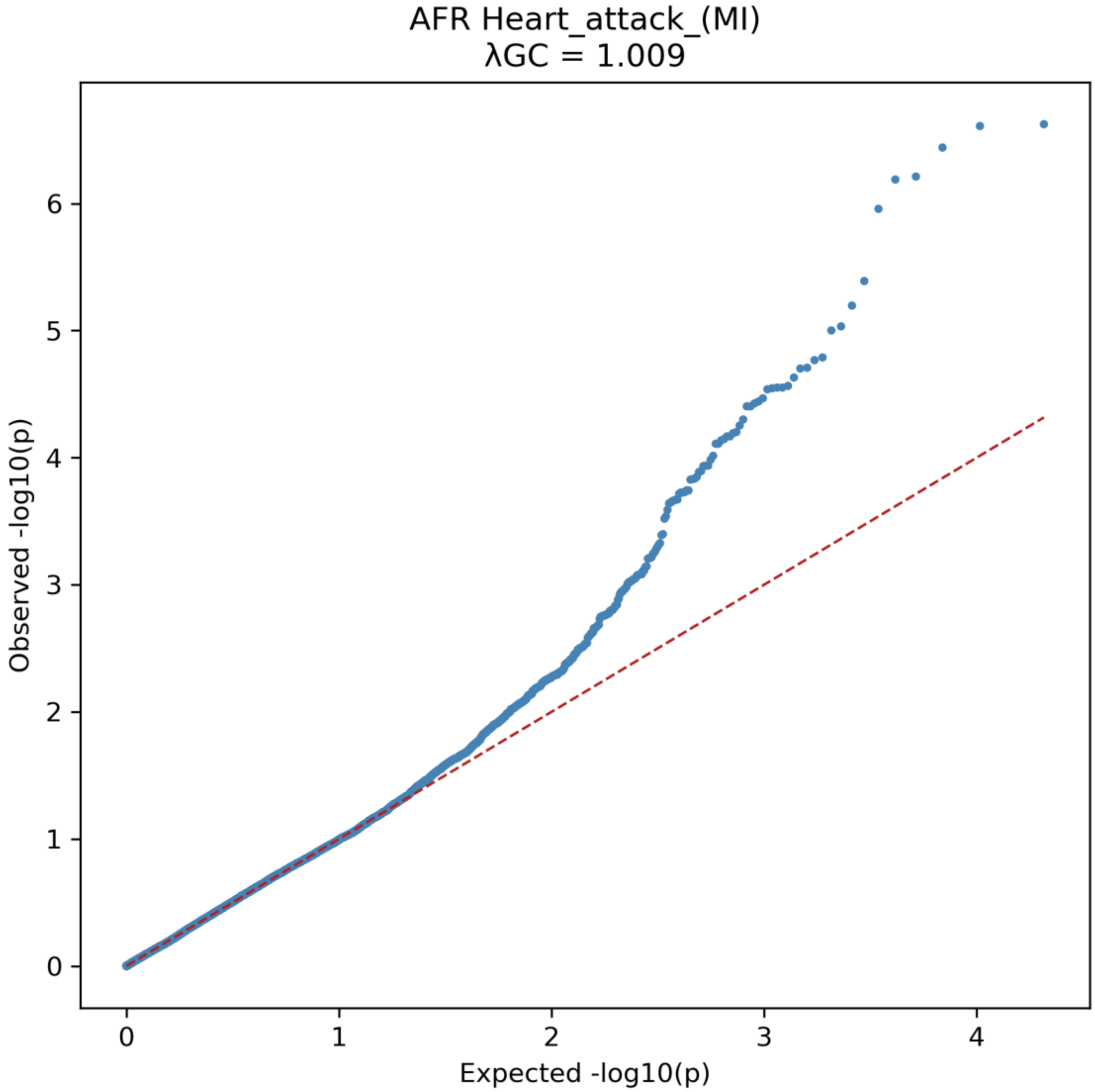


**Supplementary Figure 4**. Quantile–quantile plot for Heart attack (MI) in the AFR ancestry group. Observed versus expected −log10(p) values are shown, with λGC = 1.009. The red dashed line indicates the null expectation under no association.

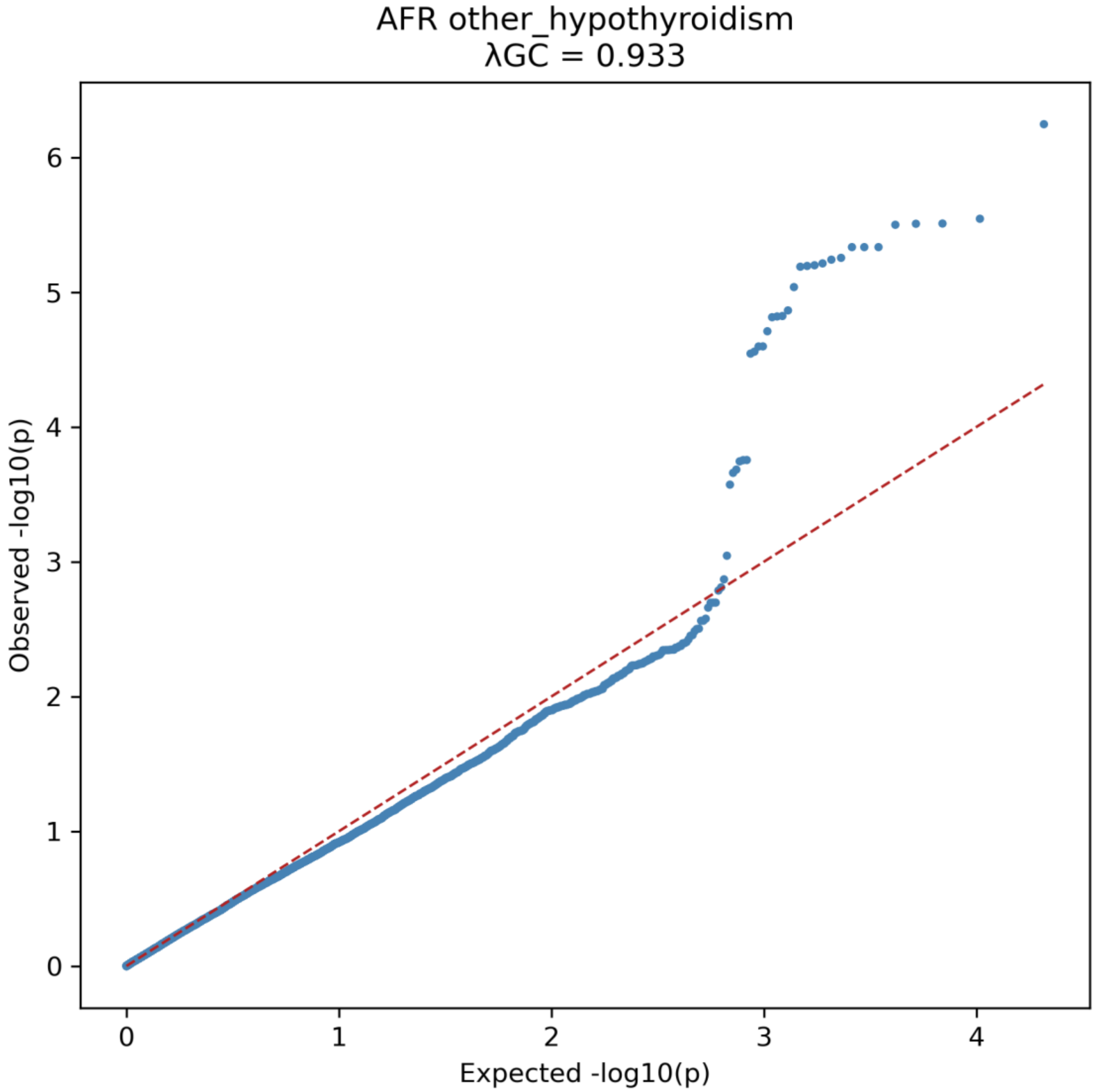


**Supplementary Figure 5**. Quantile–quantile plot for other hypothyroidism in the AFR ancestry group. Observed versus expected −log10(p) values are shown, with λGC = 0.933. The red dashed line indicates the null expectation under no association.

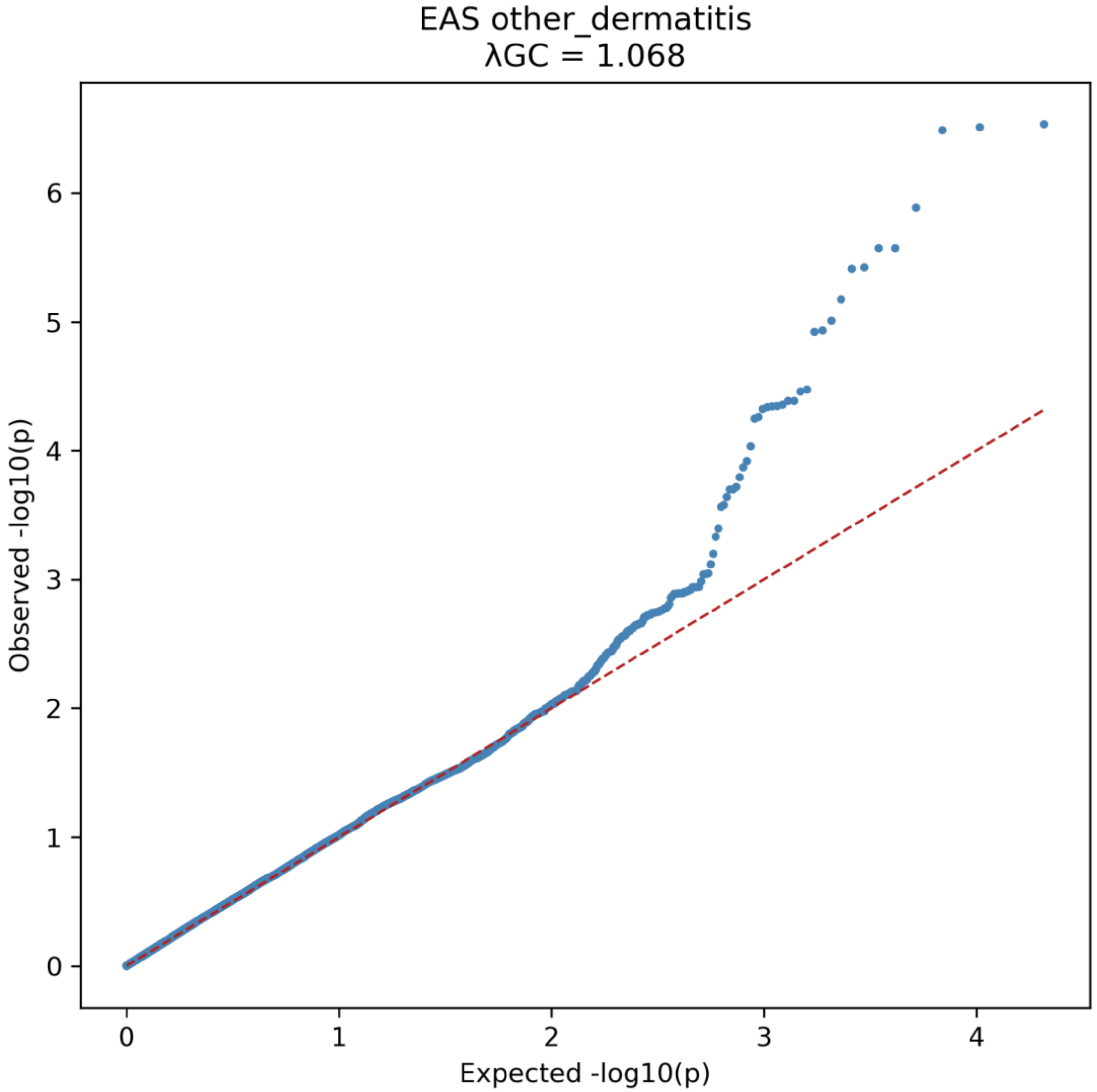


**Supplementary Figure 6**. Quantile–quantile plot for other dermatitis in the EAS ancestry group. Observed versus expected −log10(p) values are shown, with λGC = 1.068. The red dashed line indicates the null expectation under no association.

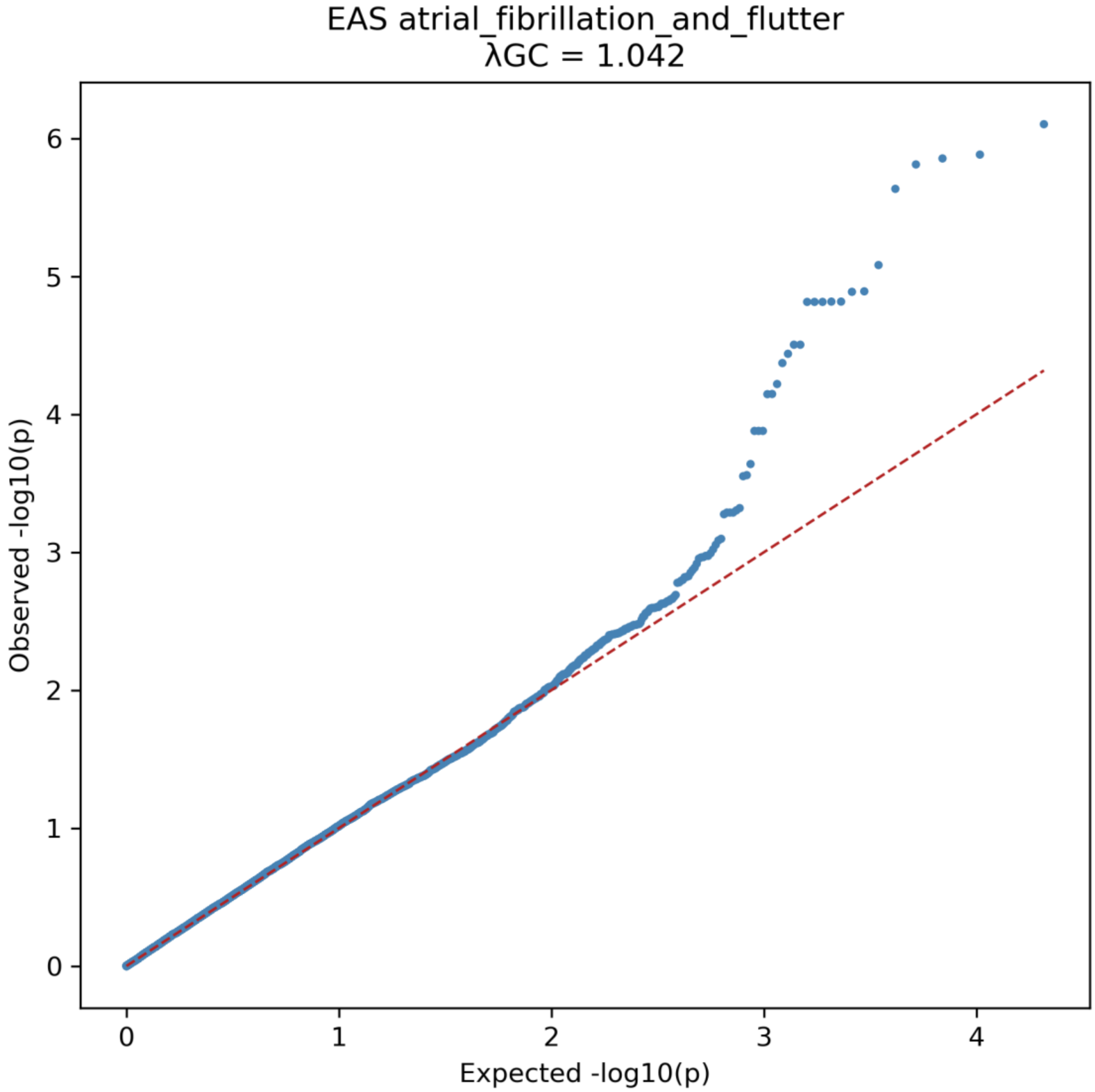


**Supplementary Figure 7**. Quantile–quantile plot for atrial fibrillation and flutter in the EAS ancestry group. Observed versus expected −log10(p) values are shown, with λGC = 1.042. The red dashed line indicates the null expectation under no association.

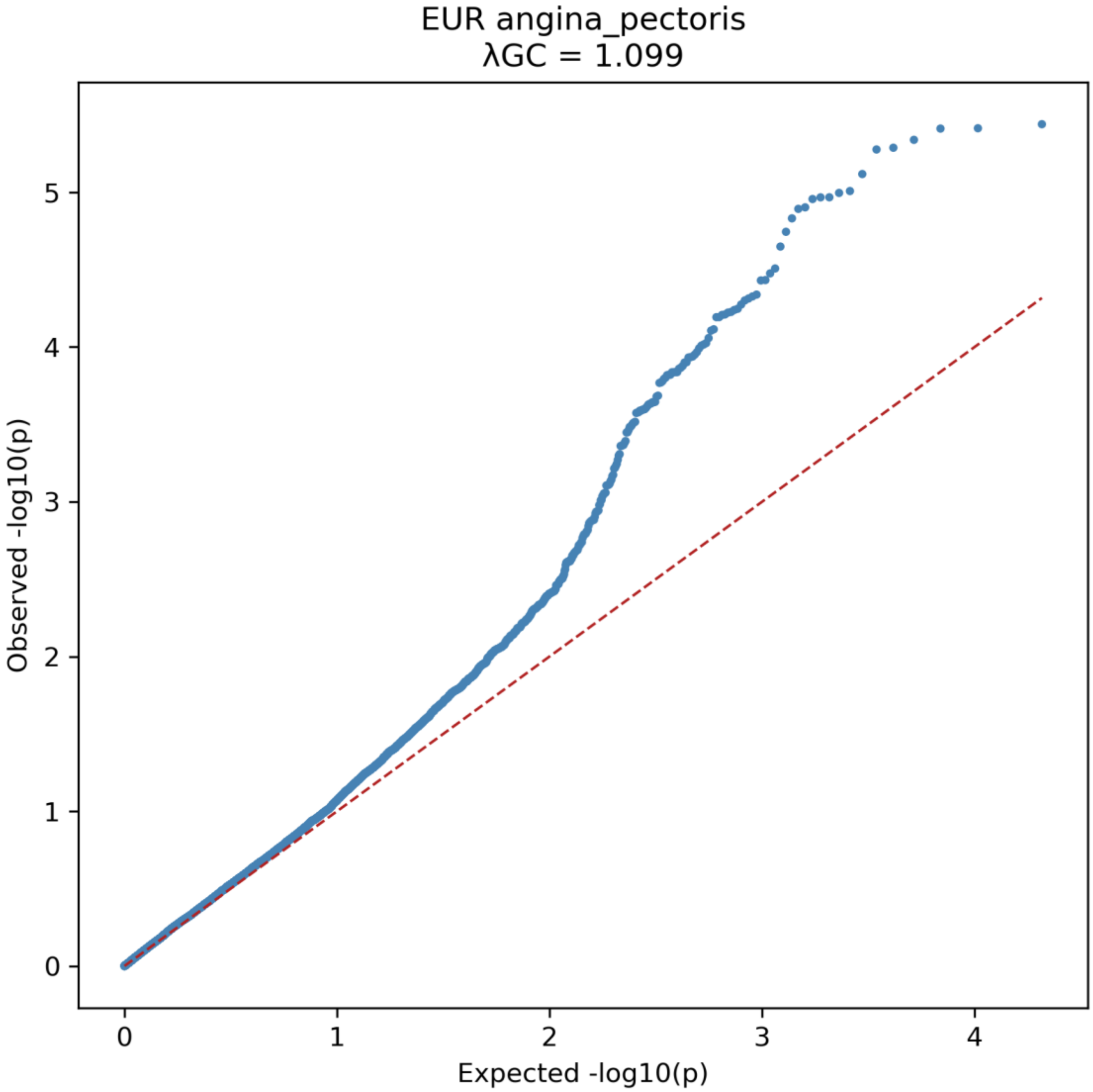


**Supplementary Figure 8**. Quantile–quantile plot for angina pectoris in the EUR ancestry group. Observed versus expected −log10(p) values are shown, with λGC = 1.099. The red dashed line indicates the null expectation under no association.

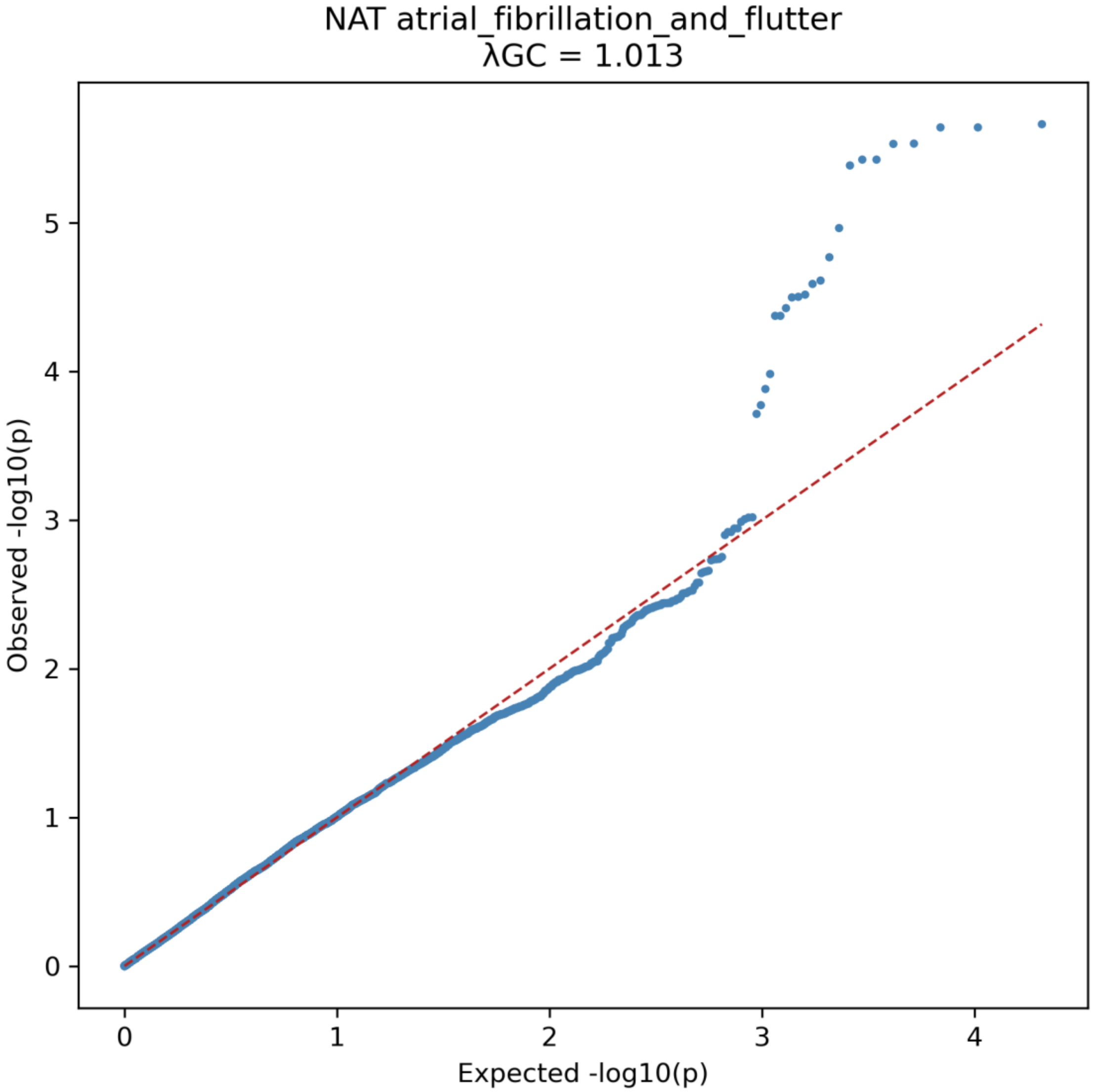


**Supplementary Figure 9**. Quantile–quantile plot for atrial fibrillation and flutter in the NAT ancestry group. Observed versus expected −log10(p) values are shown, with λGC = 1.013. The red dashed line indicates the null expectation under no association.

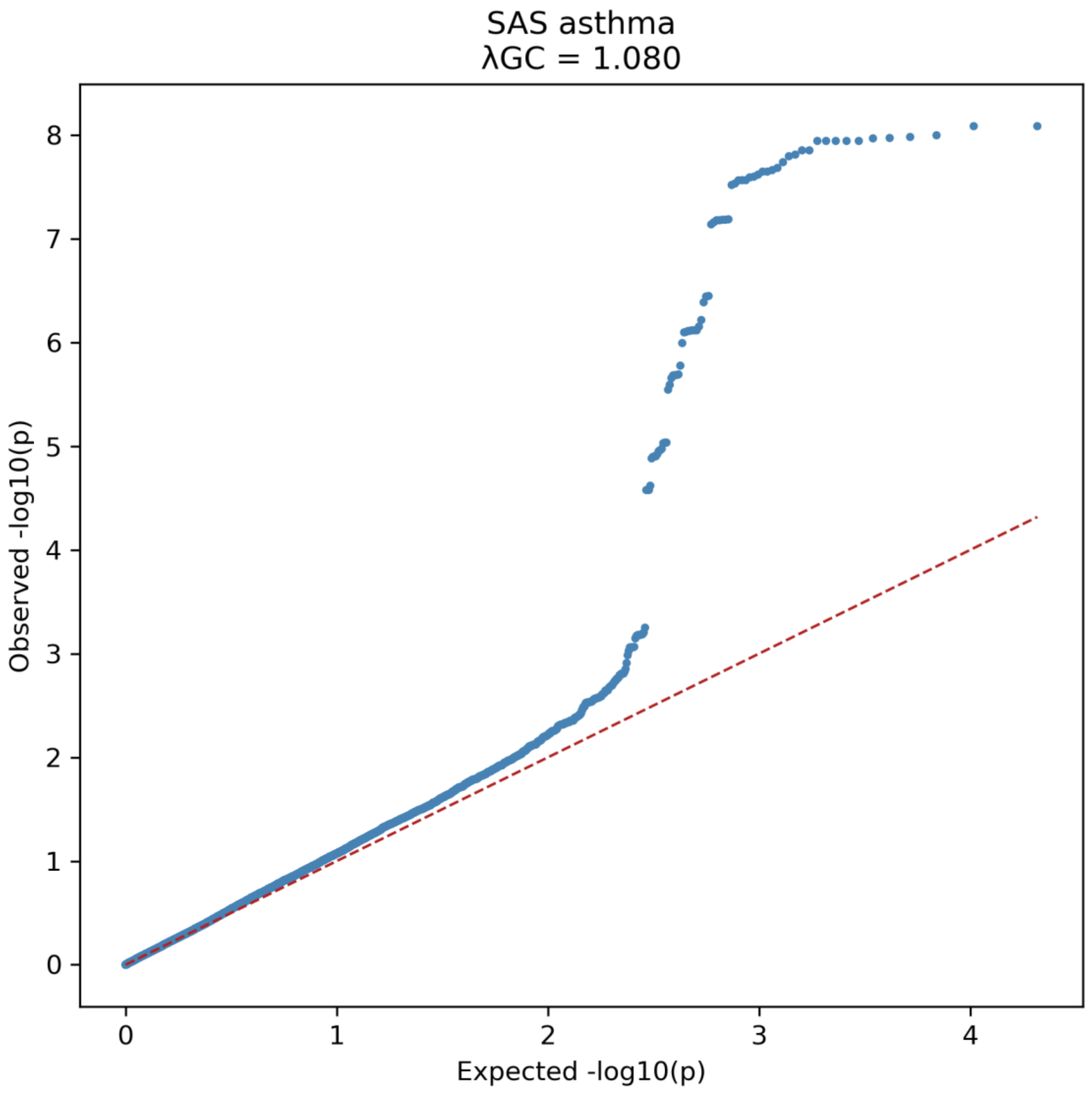


**Supplementary Figure 10**. Quantile–quantile plot for AD asthma in the SAS ancestry group. Observed versus expected −log10(p) values are shown, with λGC = 1.080. The red dashed line indicates the null expectation under no association.

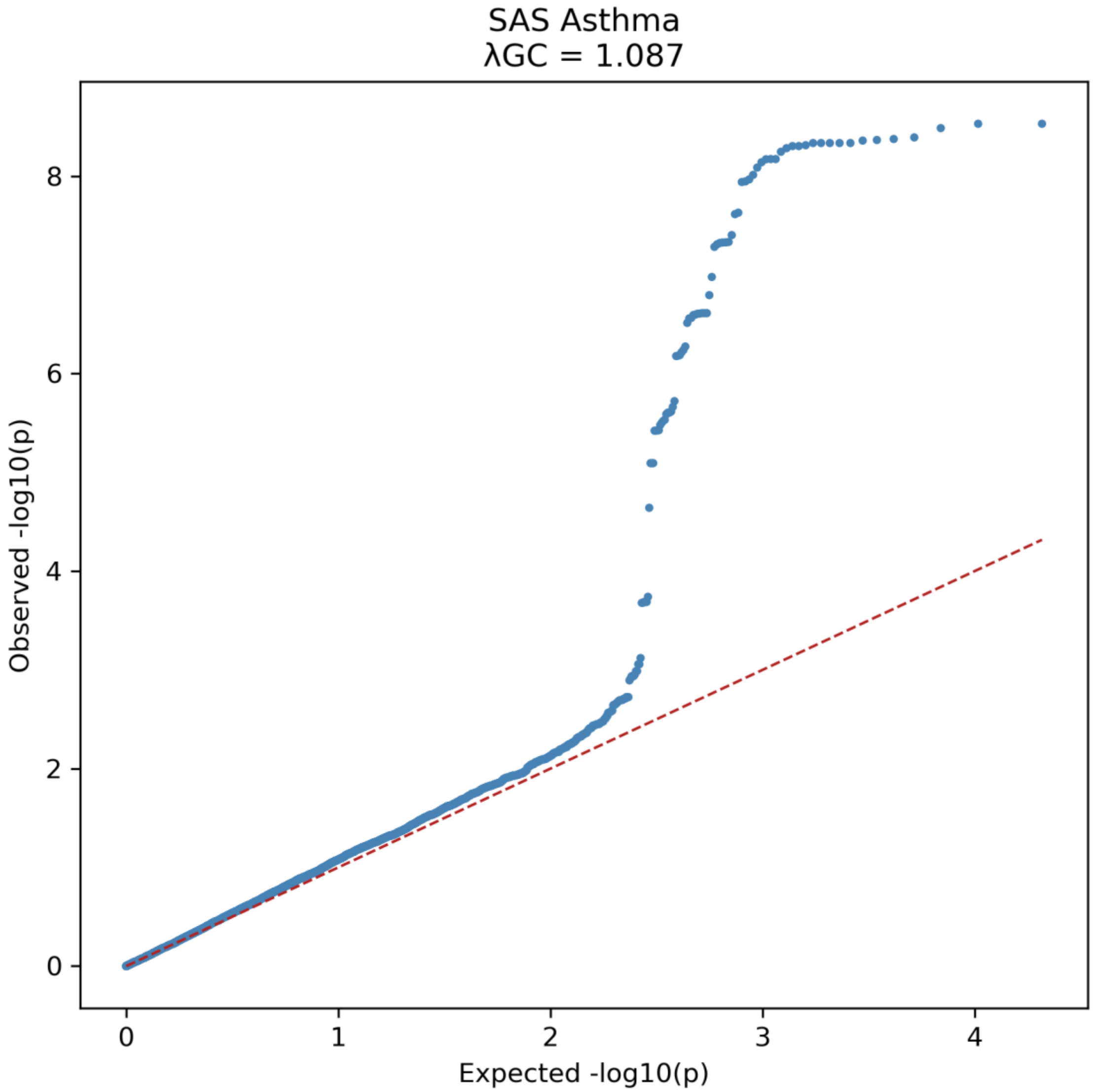


**Supplementary Figure 11**. Quantile–quantile plot for Asthma in the SAS ancestry group. Observed versus expected −log10(p) values are shown, with λGC = 1.087. The red dashed line indicates the null expectation under no association.

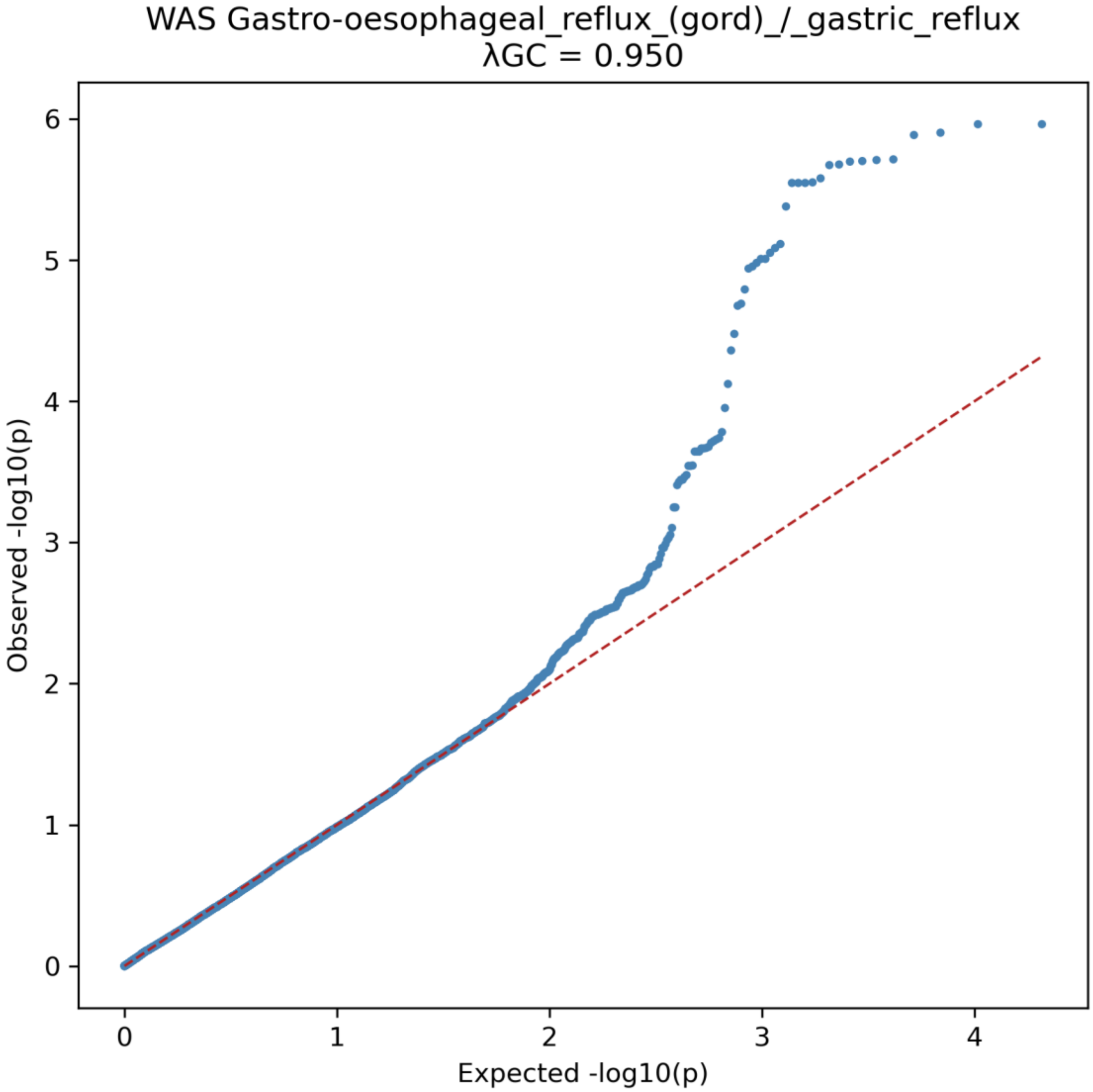


**Supplementary Figure 12**. Quantile–quantile plot for Gastro-oesophageal reflux (GORD) / gastric reflux in the WAS ancestry group. Observed versus expected −log10(p) values are shown, with λGC = 0.950. The red dashed line indicates the null expectation under no association.

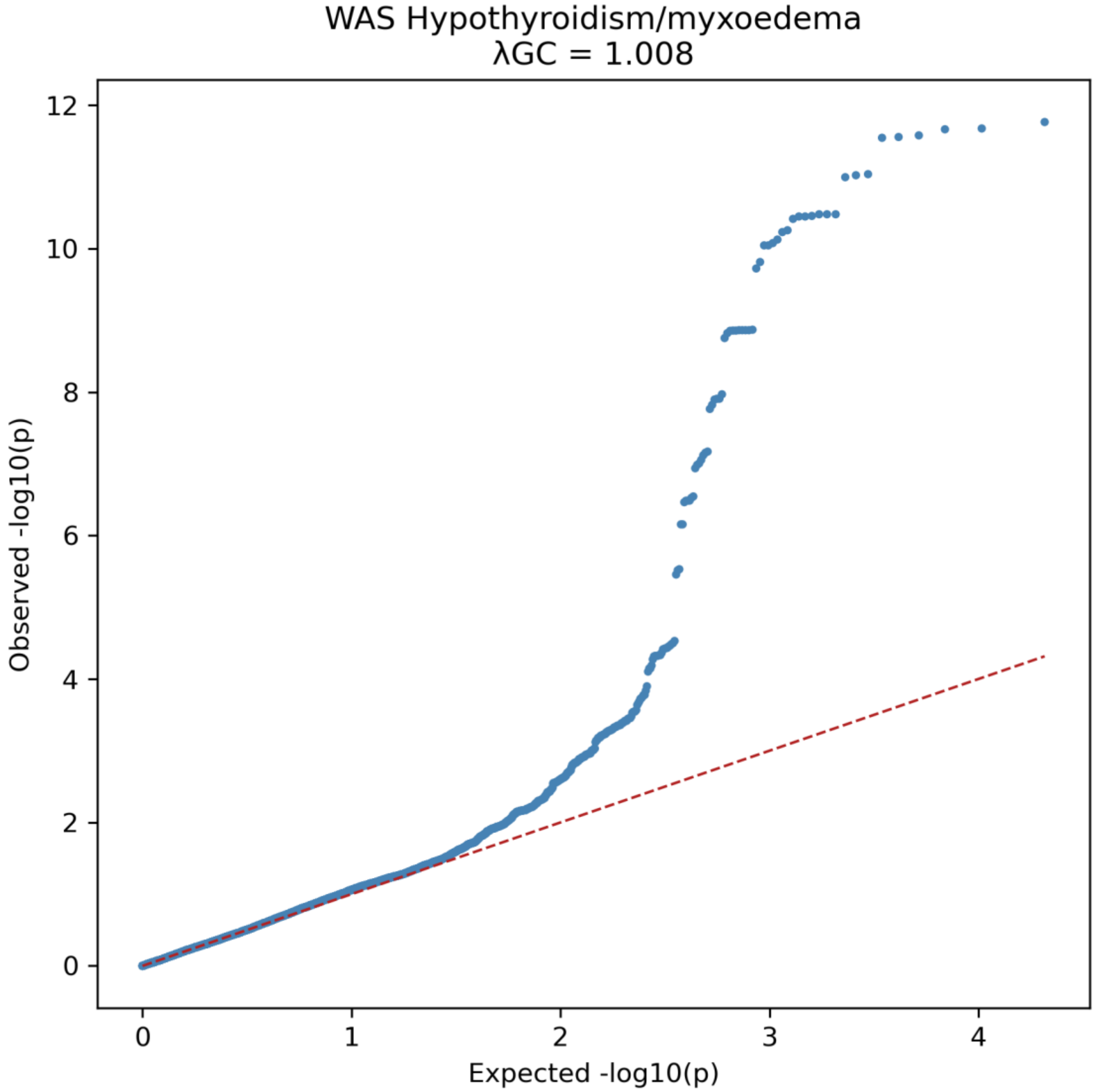


**Supplementary Figure 13**. Quantile–quantile plot for Hypothyroidism/myxoedema in the WAS ancestry group. Observed versus expected −log10(p) values are shown, with λGC = 1.008. The red dashed line indicates the null expectation under no association.

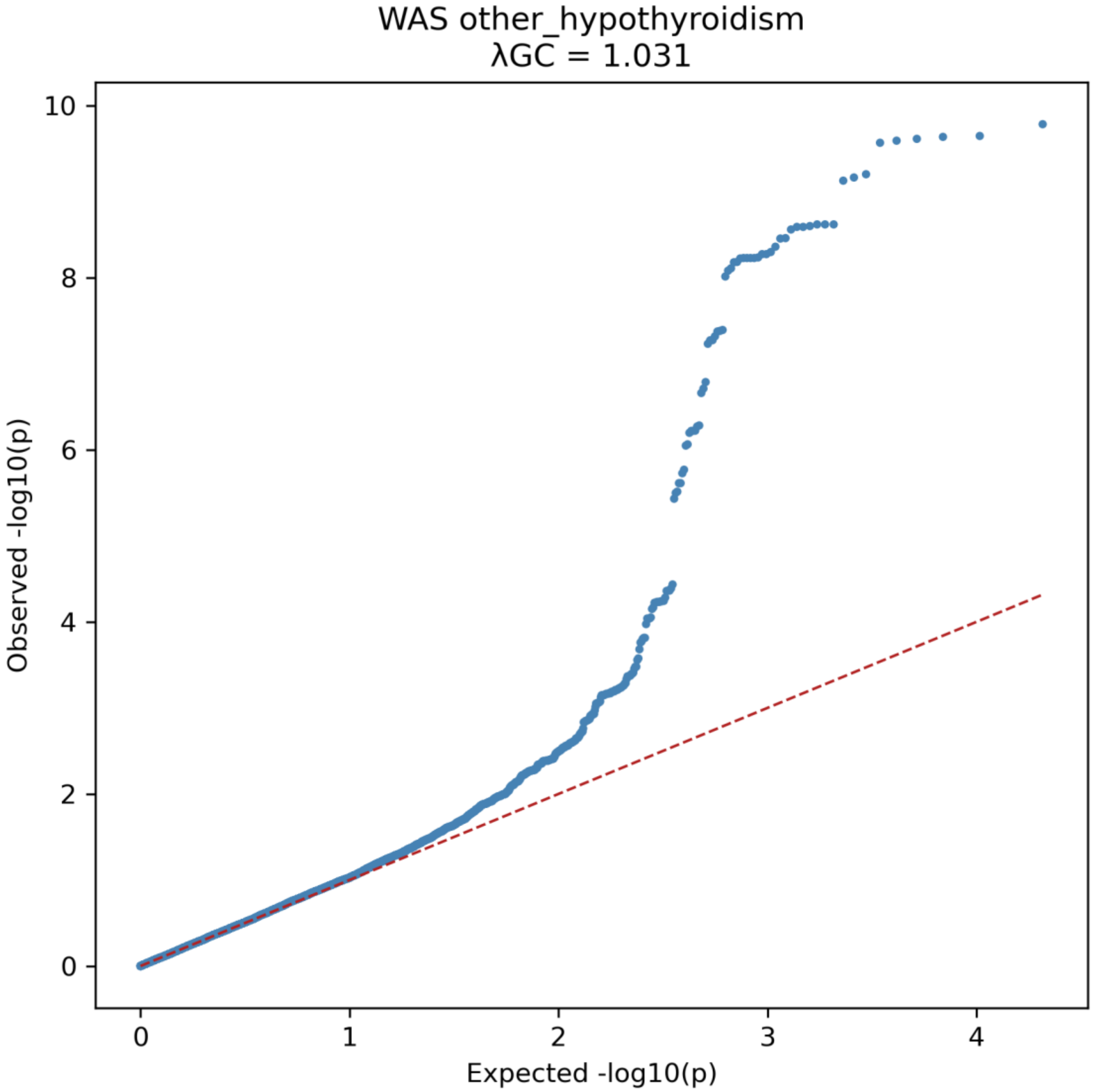


**Supplementary Figure 14**. Quantile–quantile plot for other hypothyroidism in the WAS ancestry group. Observed versus expected −log10(p) values are shown, with λGC = 1.031. The red dashed line indicates the null expectation under no association.

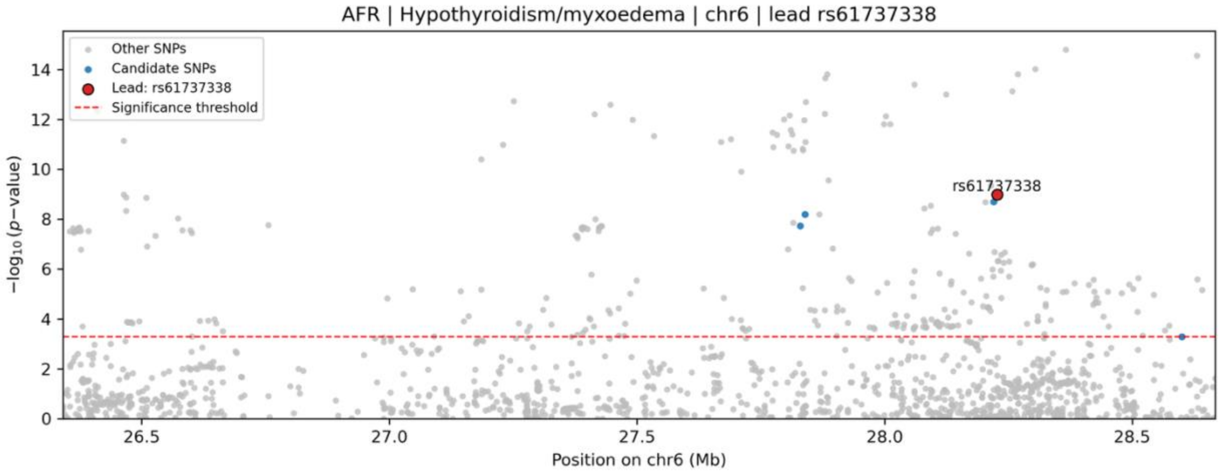


**Supplementary Figure 15**. Regional association (locus zoom) plot for Hypothyroidism/myxoedema in the AFR ancestry group at the chromosome 6 locus. Grey points denote all tested variants, blue points indicate candidate SNPs, and the red circle marks the lead variant rs61737338. The red dashed line denotes the significance threshold.

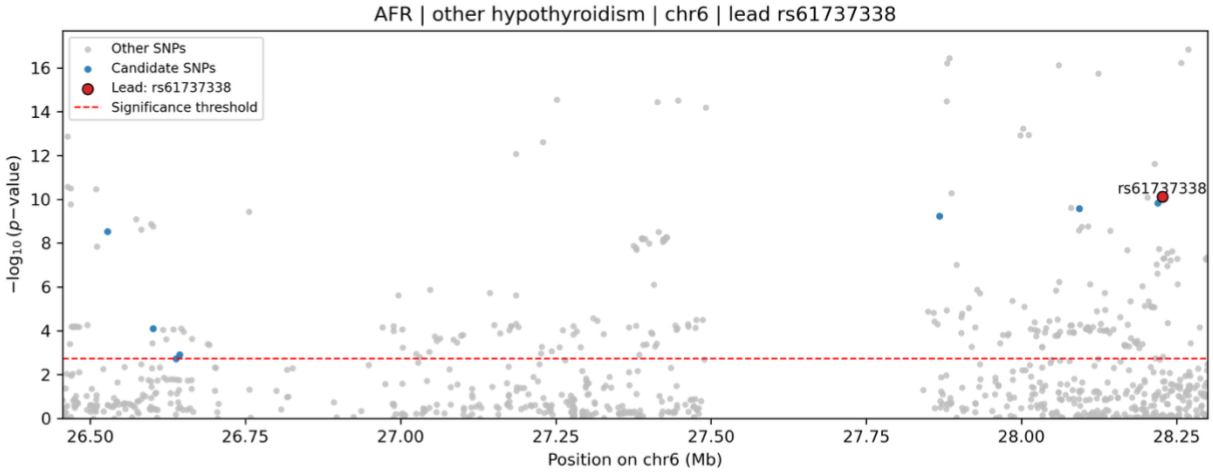


**Supplementary Figure 16**. Regional association (locus zoom) plot for other hypothyroidism in the AFR ancestry group at the chromosome 6 locus. Grey points denote all tested variants, blue points indicate candidate SNPs, and the red circle marks the lead variant rs61737338. The red dashed line denotes the significance threshold.

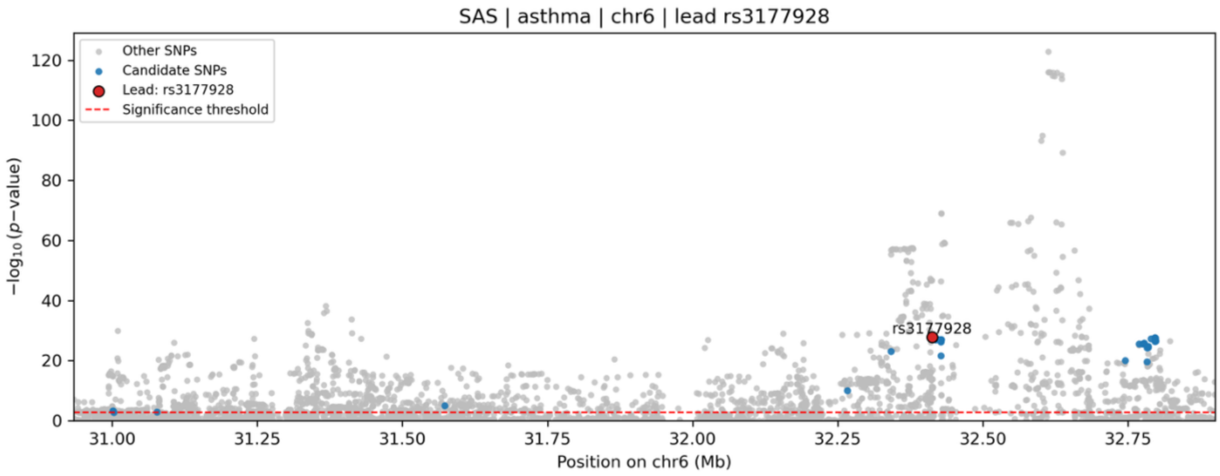


**Supplementary Figure 17**. Regional association (locus zoom) plot for AD asthma in the SAS ancestry group at the chromosome 6 locus. Grey points denote all tested variants, blue points indicate candidate SNPs, and the red circle marks the lead variant rs3177928. The red dashed line denotes the significance threshold.

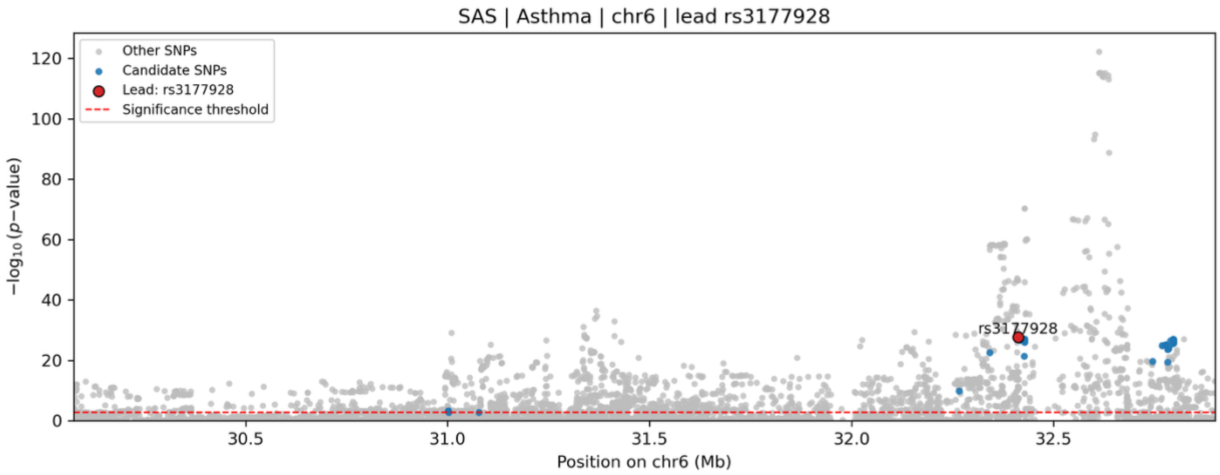


**Supplementary Figure 18**. Regional association (locus zoom) plot for Asthma in the SAS ancestry group at the chromosome 6 locus. Grey points denote all tested variants, blue points indicate candidate SNPs, and the red circle marks the lead variant rs3177928. The red dashed line denotes the significance threshold.

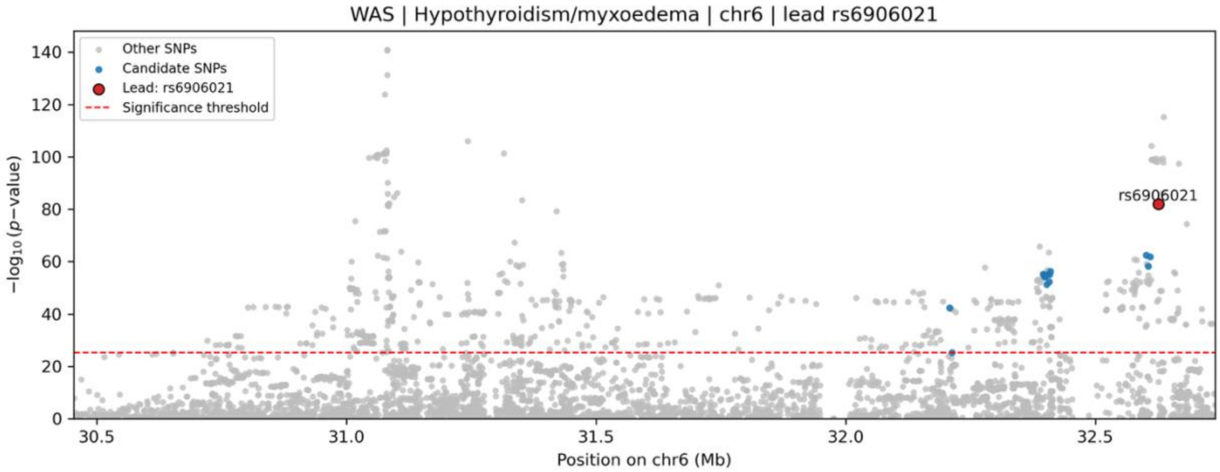


**Supplementary Figure 19**. Regional association (locus zoom) plot for Hypothyroidism/myxoedema in the WAS ancestry group at the chromosome 6 locus. Grey points denote all tested variants, blue points indicate candidate SNPs, and the red circle marks the lead variant rs6906021. The red dashed line denotes the significance threshold.

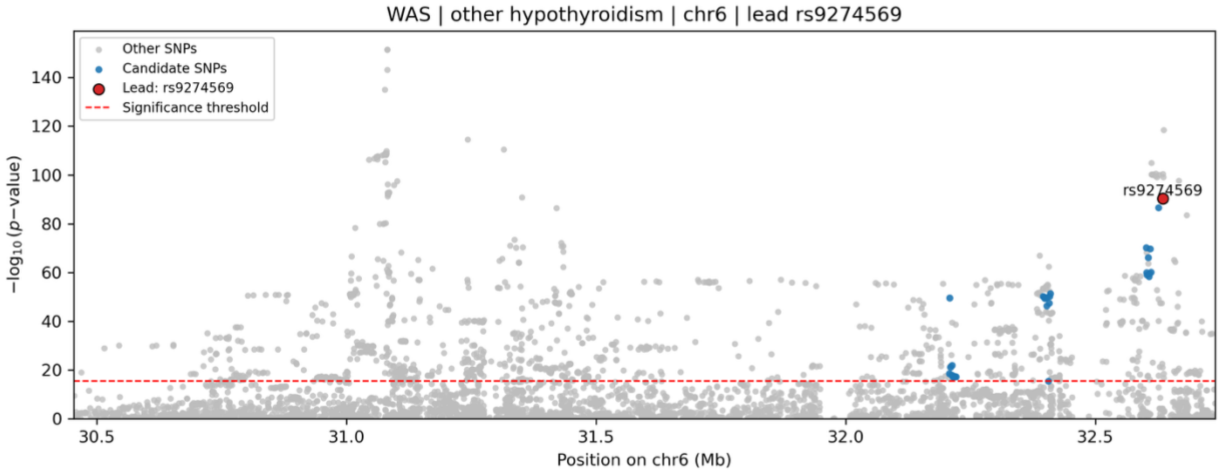


**Supplementary Figure 20**. Regional association (locus zoom) plot for other hypothyroidism in the WAS ancestry group at the chromosome 6 locus. Grey points denote all tested variants, blue points indicate candidate SNPs, and the red circle marks the lead variant rs9274569. The red dashed line denotes the significance threshold.

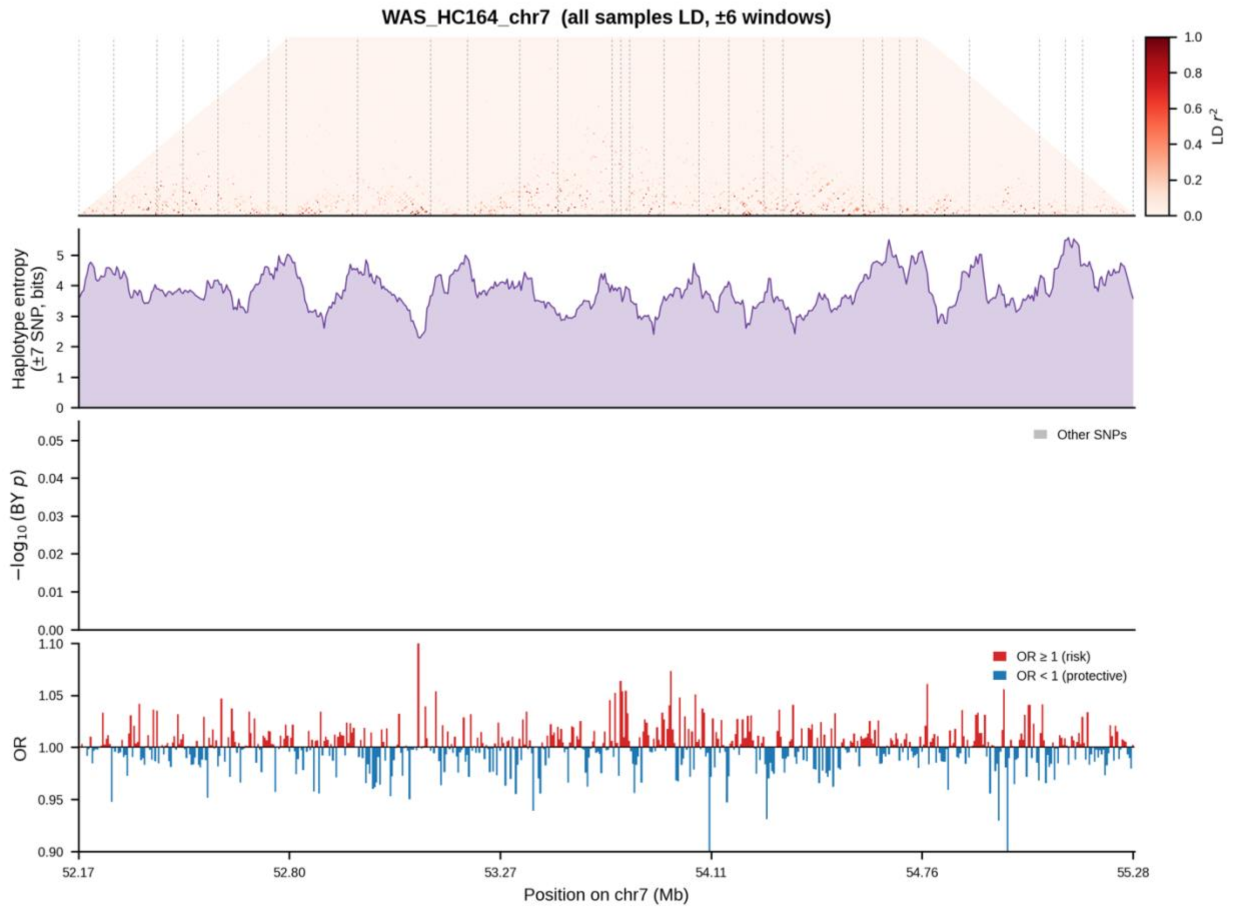


**Supplementary Figure 21**. Linkage disequilibrium, haplotype entropy, and ancestry-conditioned association signal for Gastro-oesophageal reflux (GORD) / gastric reflux in the WAS ancestry group at the chromosome 7 locus. From top to bottom: pairwise LD (r²) computed across all samples (predominantly EUR-like ancestry); local haplotype entropy (±6-SNP window, bits); BY-corrected −log10(p) values from the ancestry-associated conditional model, with the fine-mapped candidate SNP highlighted in blue; and per-SNP odds ratios, colored by direction of effect (red, OR ≥ 1; blue, OR < 1).

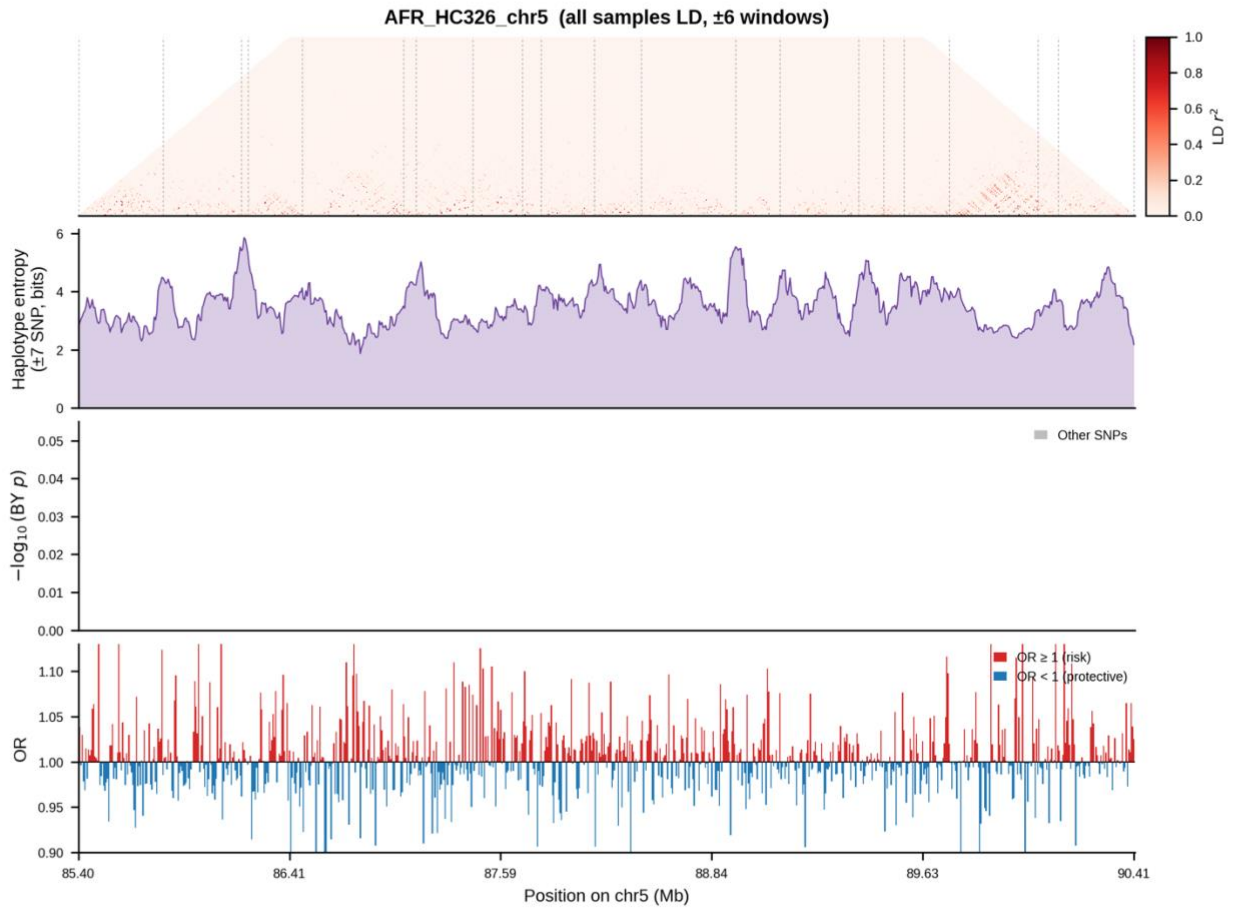


**Supplementary Figure 22**. Linkage disequilibrium, haplotype entropy, and ancestry-conditioned association signal for Heart attack (MI) in the AFR ancestry group at the chromosome 5 locus. From top to bottom: pairwise LD ($r^2$) computed across all samples (predominantly EUR-like ancestry); local haplotype entropy (±6-SNP window, bits); BY-corrected $-\log10(p)$ values from the ancestry-associated conditional model, with the fine-mapped candidate SNP highlighted in blue; and per-SNP odds ratios, colored by direction of effect (red, OR ≥ 1; blue, OR < 1).

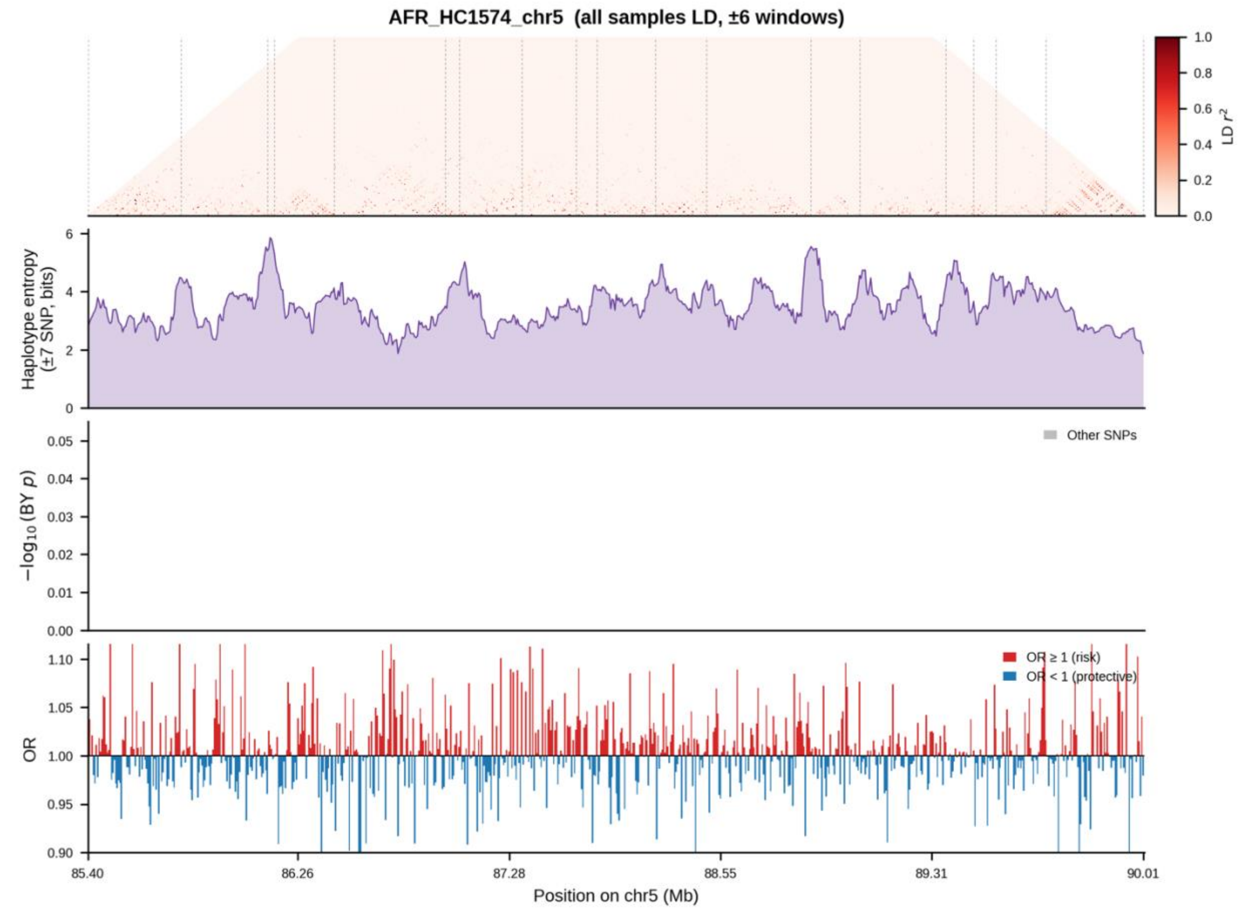


**Supplementary Figure 23**. Linkage disequilibrium, haplotype entropy, and ancestry-conditioned association signal for AD myocardial infarction in the AFR ancestry group at the chromosome 5 locus. From top to bottom: pairwise LD ($r^2$) computed across all samples (predominantly EUR-like ancestry); local haplotype entropy (±6-SNP window, bits); BY-corrected $-\log10(p)$ values from the ancestry-associated conditional model, with the fine-mapped candidate SNP highlighted in blue; and per-SNP odds ratios, colored by direction of effect (red, OR ≥ 1; blue, OR < 1).

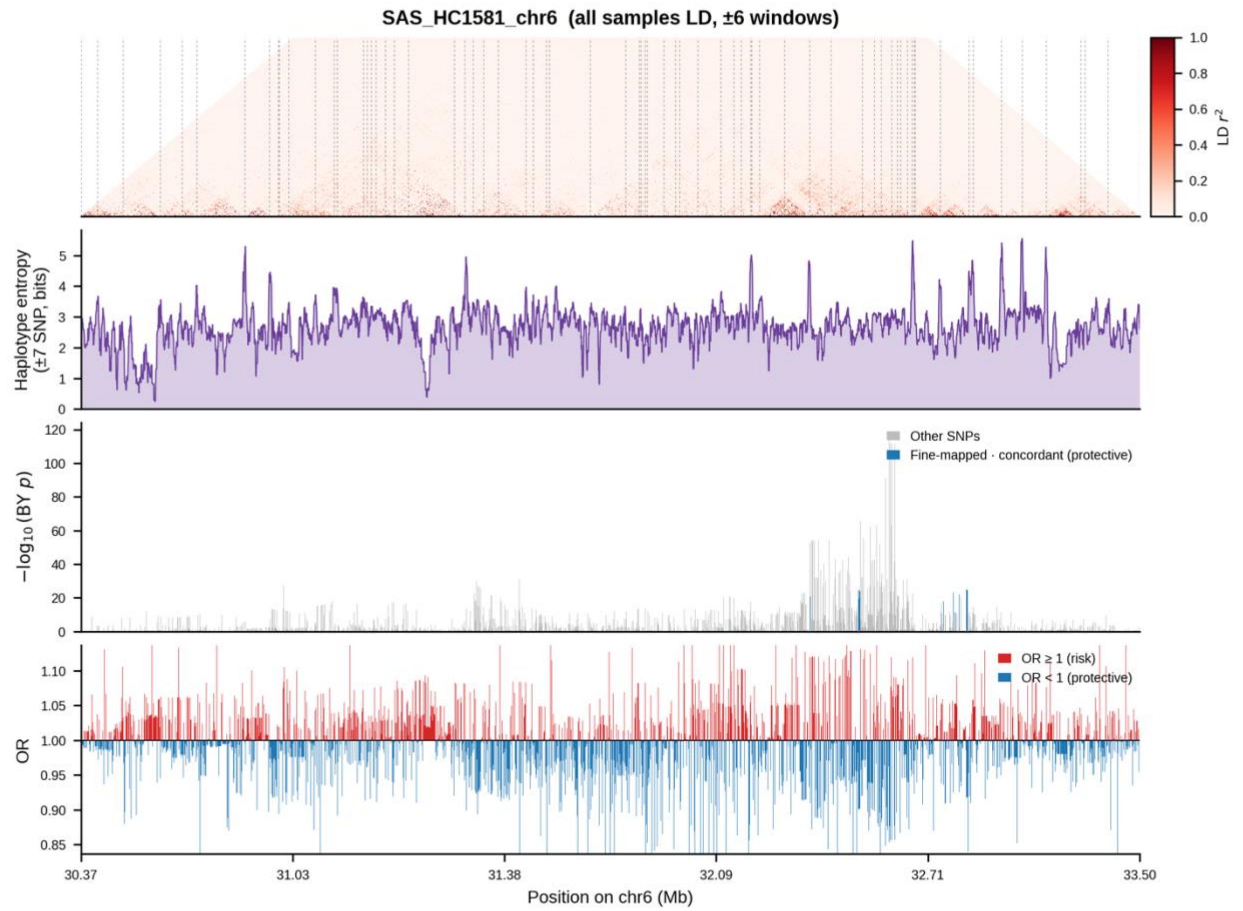


**Supplementary Figure 24**. Linkage disequilibrium, haplotype entropy, and ancestry-conditioned association signal for AD asthma in the SAS ancestry group at the chromosome 6 locus. From top to bottom: pairwise LD ($r^2$) computed across all samples (predominantly EUR-like ancestry); local haplotype entropy (±6-SNP window, bits); BY-corrected −log10(p) values from the ancestry-associated conditional model, with the fine-mapped candidate SNP highlighted in blue; and per-SNP odds ratios, colored by direction of effect (red, OR ≥ 1; blue, OR < 1).

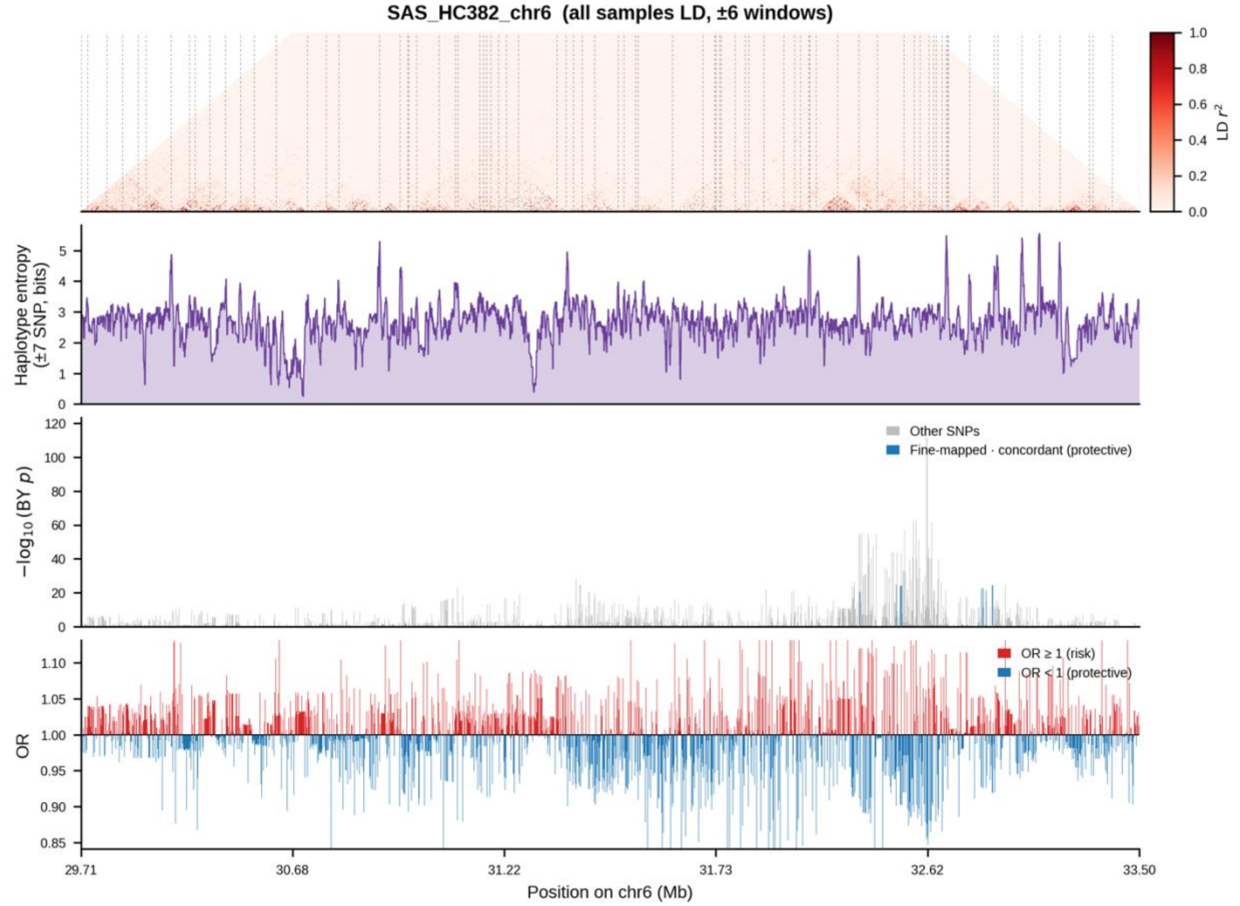


**Supplementary Figure 25**. Linkage disequilibrium, haplotype entropy, and ancestry-conditioned association signal for Asthma in the SAS ancestry group at the chromosome 6 locus. From top to bottom: pairwise LD ($r^2$) computed across all samples (predominantly EUR-like ancestry); local haplotype entropy (±6-SNP window, bits); BY-corrected $-\log10(p)$ values from the ancestry-associated conditional model, with the fine-mapped candidate SNP highlighted in blue; and per-SNP odds ratios, colored by direction of effect (red, OR ≥ 1; blue, OR < 1).

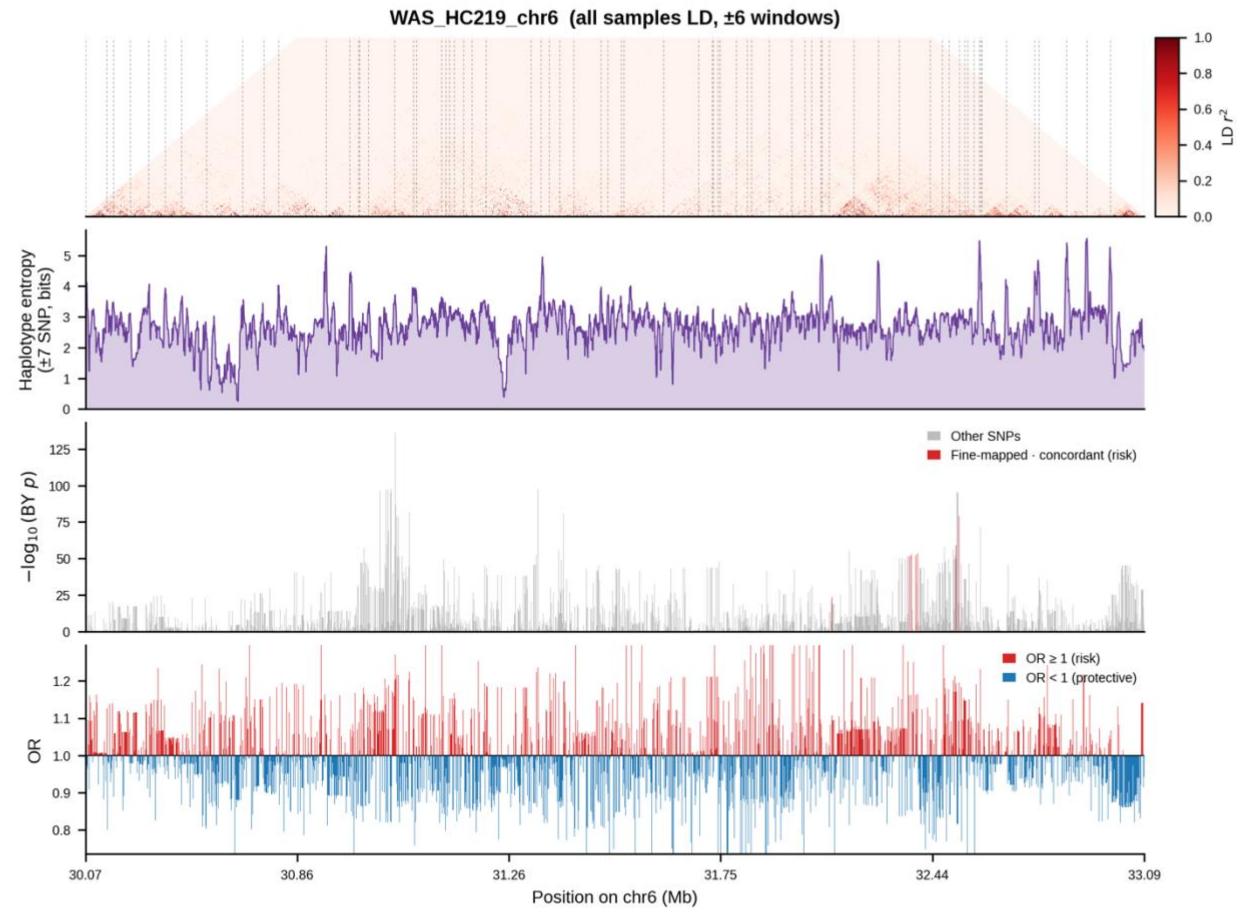


**Supplementary Figure 26**. Linkage disequilibrium, haplotype entropy, and ancestry-conditioned association signal for Hypothyroidism/myxoedema in the WAS ancestry group at the chromosome 6 locus. From top to bottom: pairwise LD (r²) computed across all samples (predominantly EUR-like ancestry); local haplotype entropy (±6-SNP window, bits); BY-corrected −log10(p) values from the ancestry-associated conditional model, with the fine-mapped candidate SNP highlighted in blue; and per-SNP odds ratios, colored by direction of effect (red, OR ≥ 1; blue, OR < 1).

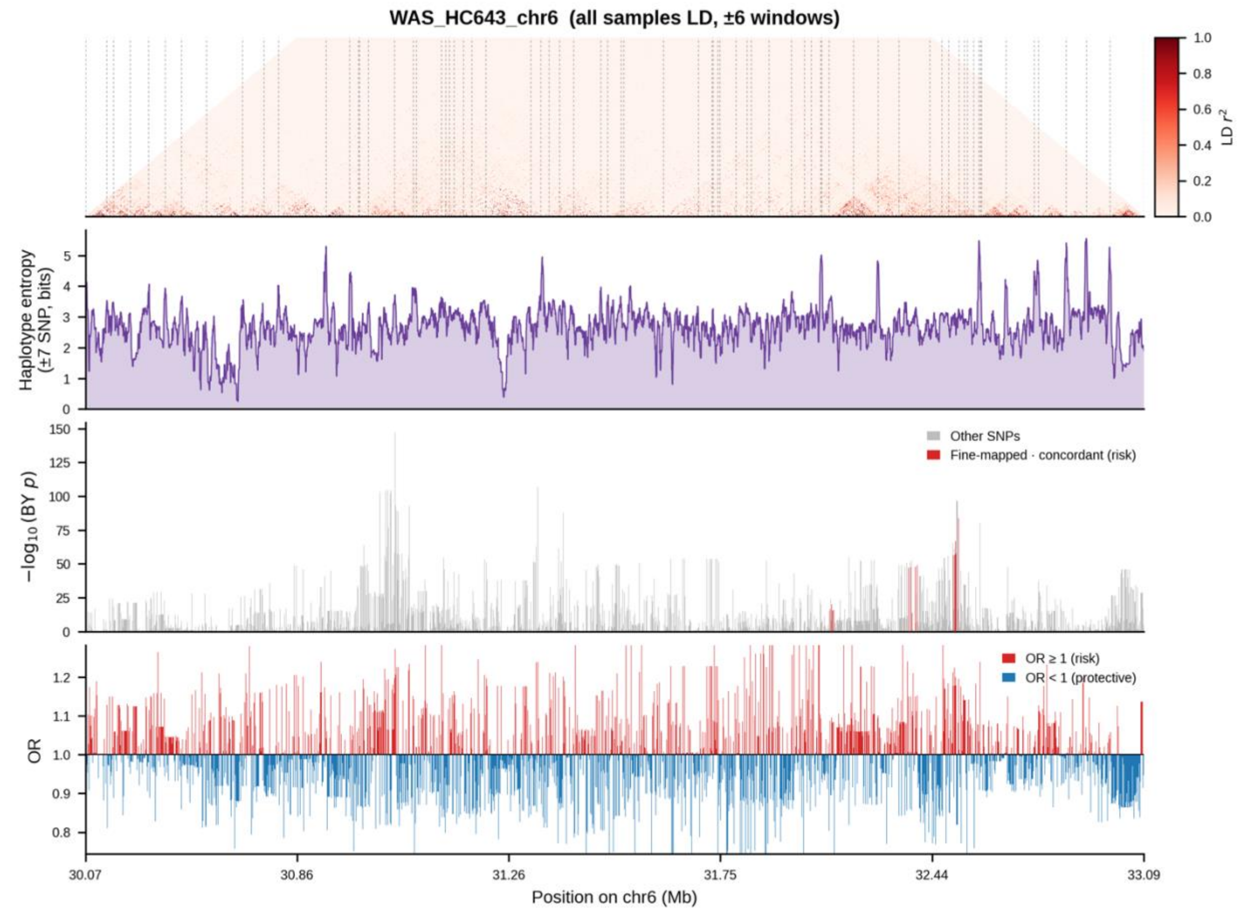


**Supplementary Figure 27**. Linkage disequilibrium, haplotype entropy, and ancestry-conditioned association signal for other hypothyroidism in the WAS ancestry group at the chromosome 6 locus. From top to bottom: pairwise LD ($r^2$) computed across all samples (predominantly EUR-like ancestry); local haplotype entropy (±6-SNP window, bits); BY-corrected $-\log10(p)$ values from the ancestry-associated conditional model, with the fine-mapped candidate SNP highlighted in blue; and per-SNP odds ratios, colored by direction of effect (red, OR ≥ 1; blue, OR < 1).

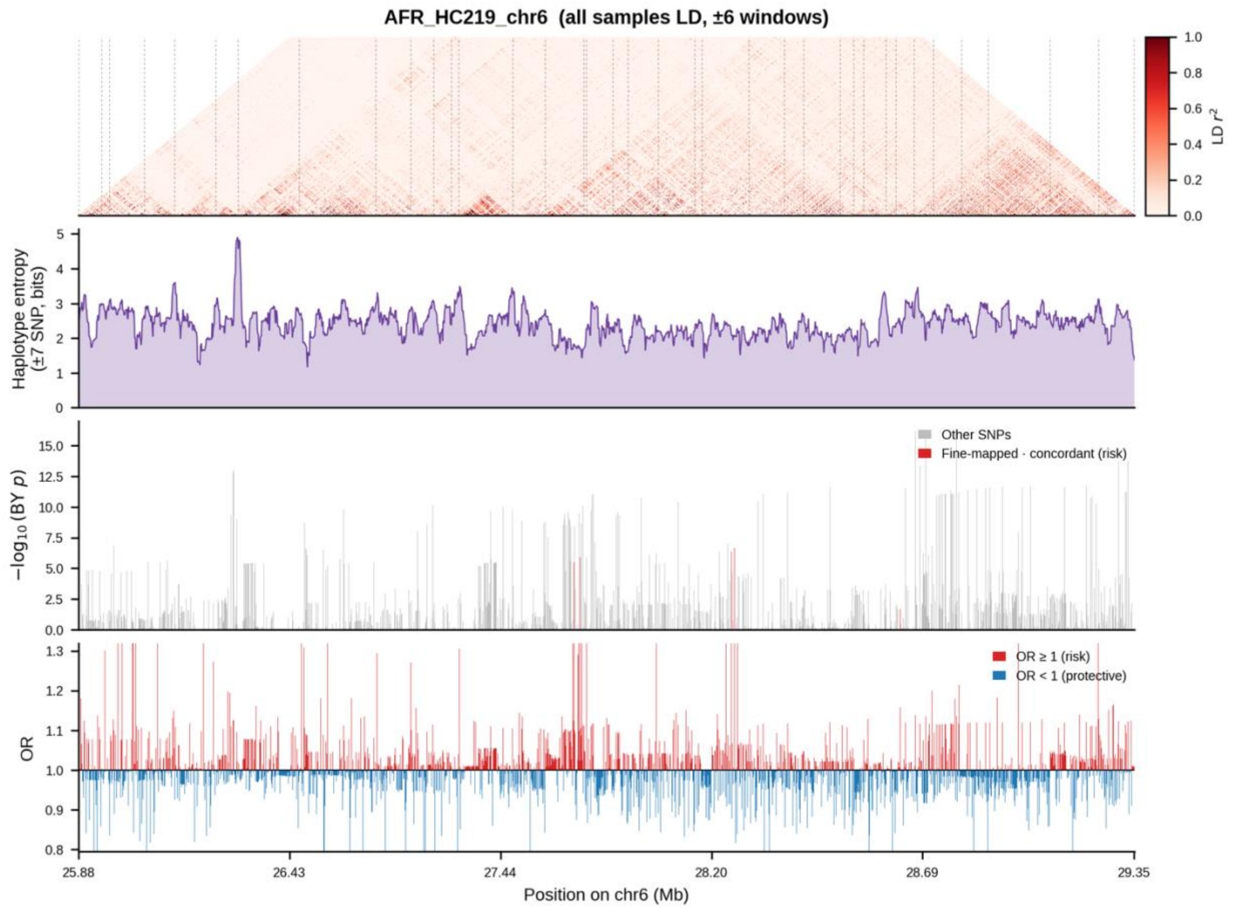


**Supplementary Figure 28**. Linkage disequilibrium, haplotype entropy, and ancestry-conditioned association signal for Hypothyroidism/myxoedema in the AFR ancestry group at the chromosome 6 locus. From top to bottom: pairwise LD ($r^2$) computed across all samples (predominantly EUR-like ancestry); local haplotype entropy (±6-SNP window, bits); BY-corrected $-\log10(p)$ values from the ancestry-associated conditional model, with the fine-mapped candidate SNP highlighted in blue; and per-SNP odds ratios, colored by direction of effect (red, OR ≥ 1; blue, OR < 1).

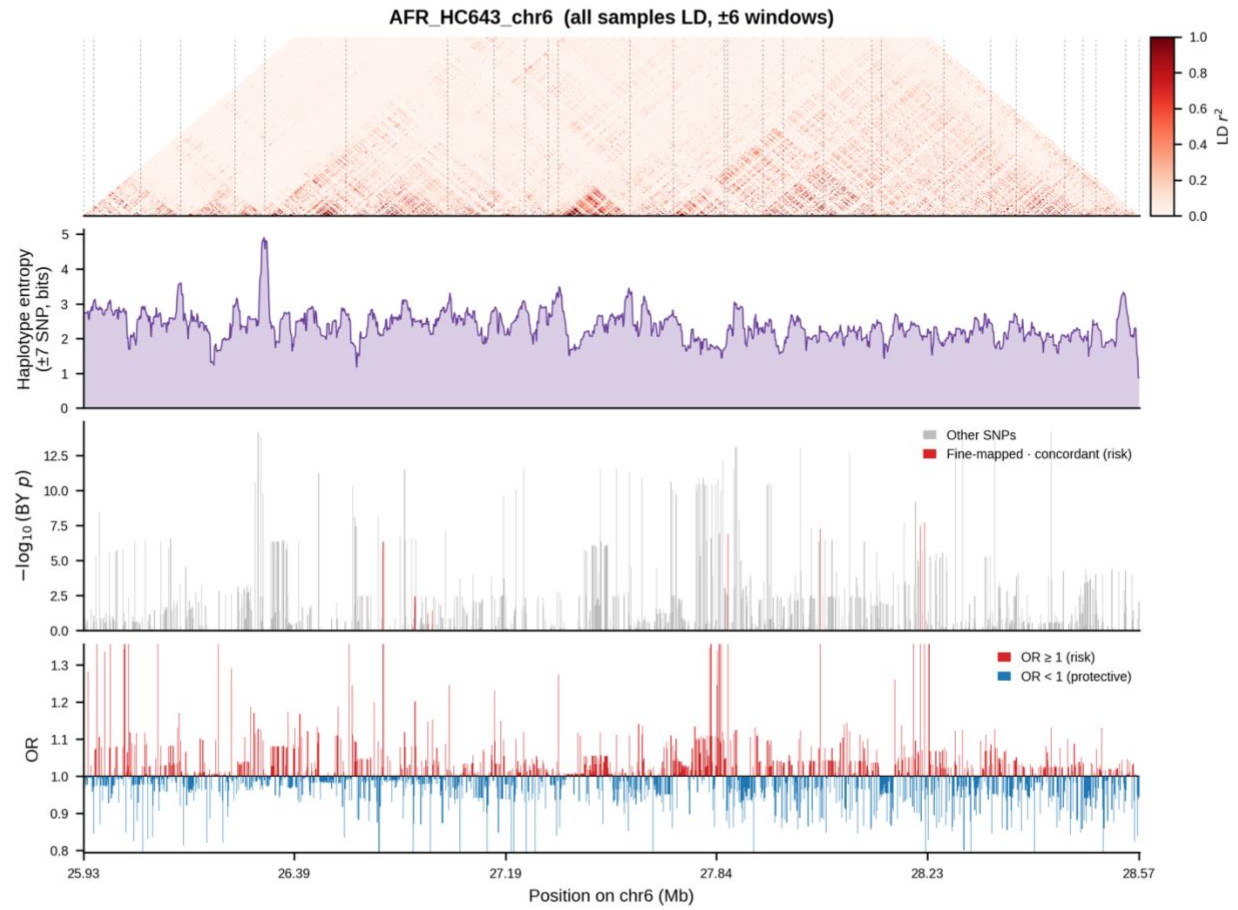


**Supplementary Figure 29**. Linkage disequilibrium, haplotype entropy, and ancestry-conditioned association signal for other hypothyroidism in the AFR ancestry group at the chromosome 6 locus. From top to bottom: pairwise LD (r²) computed across all samples (predominantly EUR-like ancestry); local haplotype entropy (±6-SNP window, bits); BY-corrected −log10(p) values from the ancestry-associated conditional model, with the fine-mapped candidate SNP highlighted in blue; and per-SNP odds ratios, colored by direction of effect (red, OR ≥ 1; blue, OR < 1).

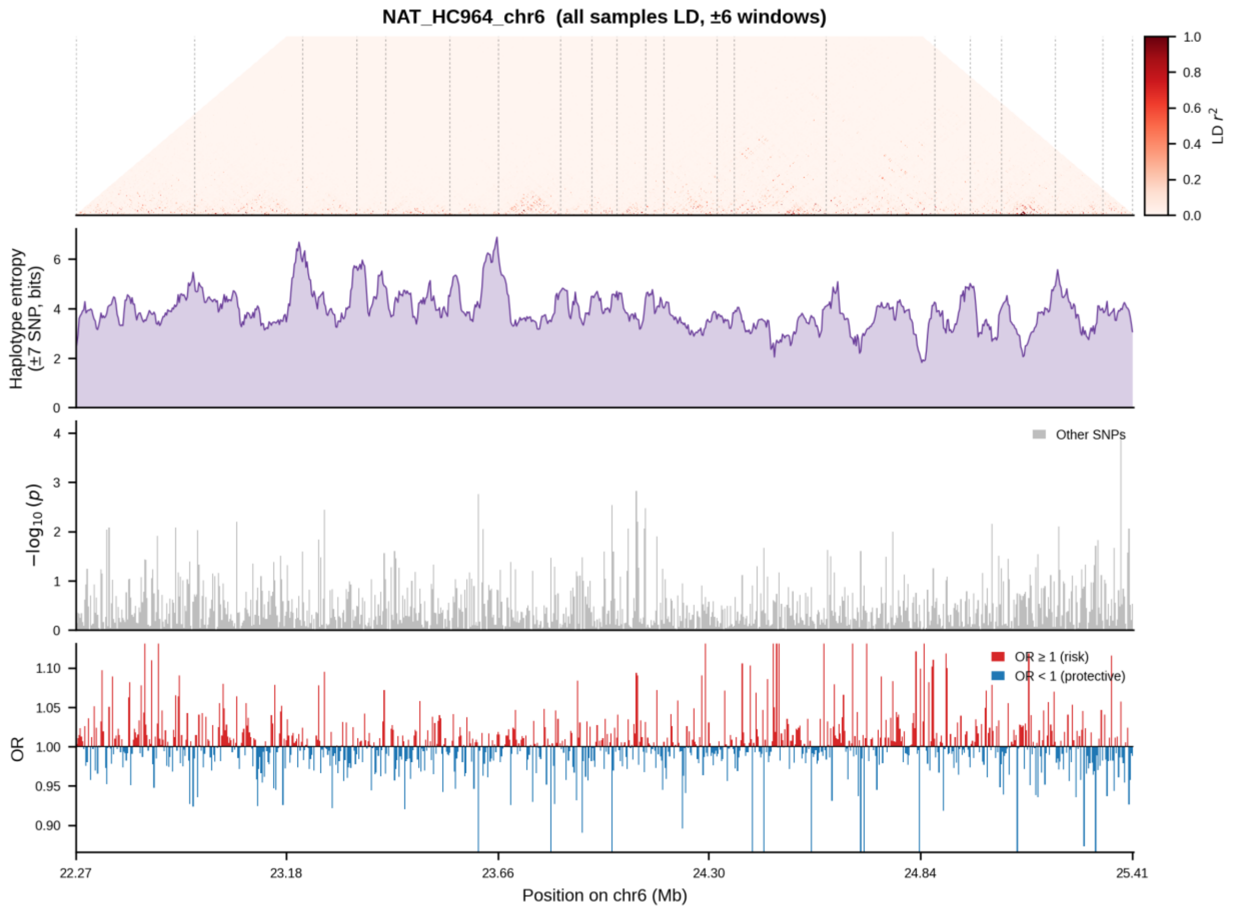


**Supplementary Figure 30**. Linkage disequilibrium, haplotype entropy, and ancestry-conditioned association signal for atrial fibrillation and flutter in the NAT ancestry group at the chromosome 6 locus. From top to bottom: pairwise LD ($r^2$) computed across all samples (predominantly EUR-like ancestry); local haplotype entropy (±6-SNP window, bits); BY-corrected $-\log10(p)$ values from the ancestry-associated conditional model, with the fine-mapped candidate SNP highlighted in blue; and per-SNP odds ratios, colored by direction of effect (red, OR ≥ 1; blue, OR < 1).

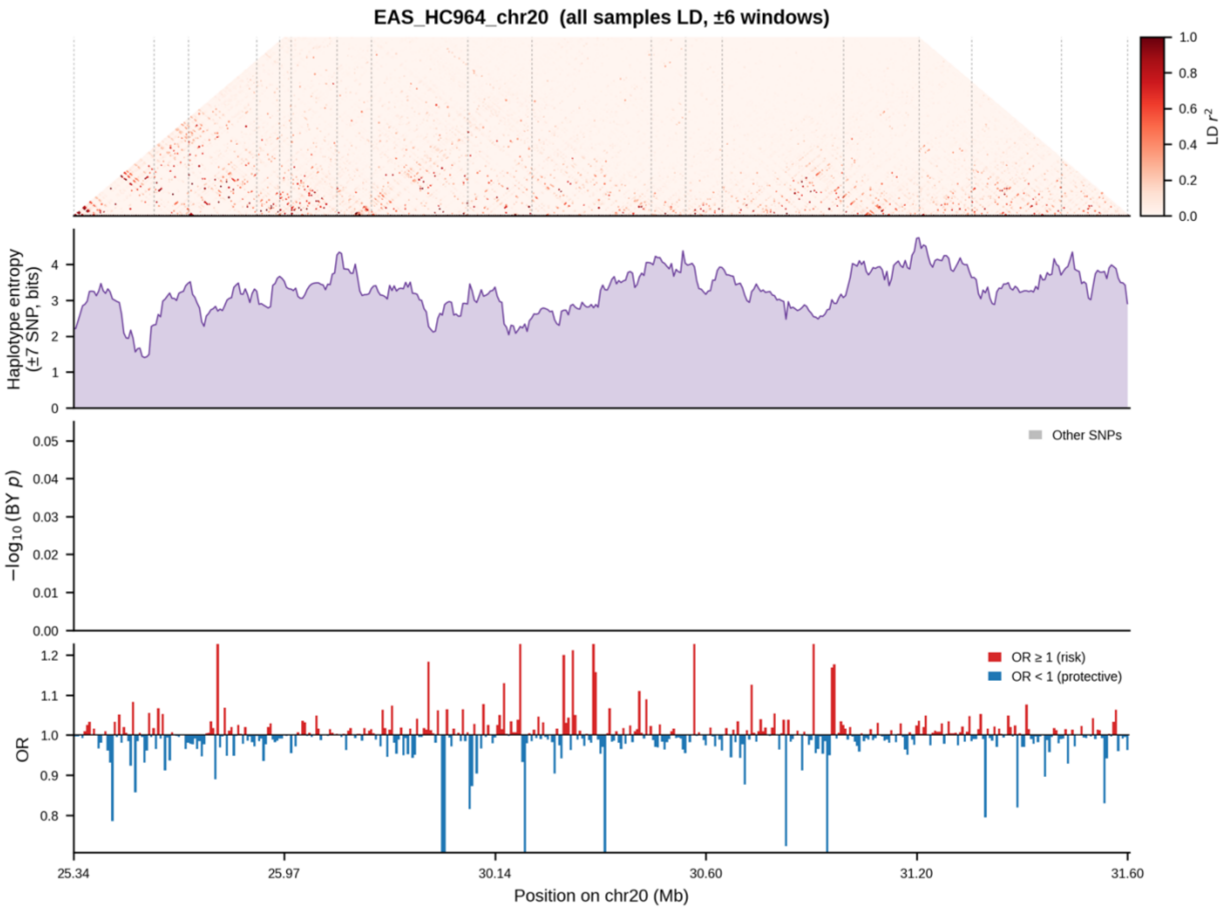


**Supplementary Figure 31**. Linkage disequilibrium, haplotype entropy, and ancestry-conditioned association signal for atrial fibrillation and flutter in the EAS ancestry group at the chromosome 20 locus. From top to bottom: pairwise LD ($r^2$) computed across all samples (predominantly EUR-like ancestry); local haplotype entropy (±6-SNP window, bits); BY-corrected −log10(p) values from the ancestry-associated conditional model, with the fine-mapped candidate SNP highlighted in blue; and per-SNP odds ratios, colored by direction of effect (red, OR ≥ 1; blue, OR < 1).

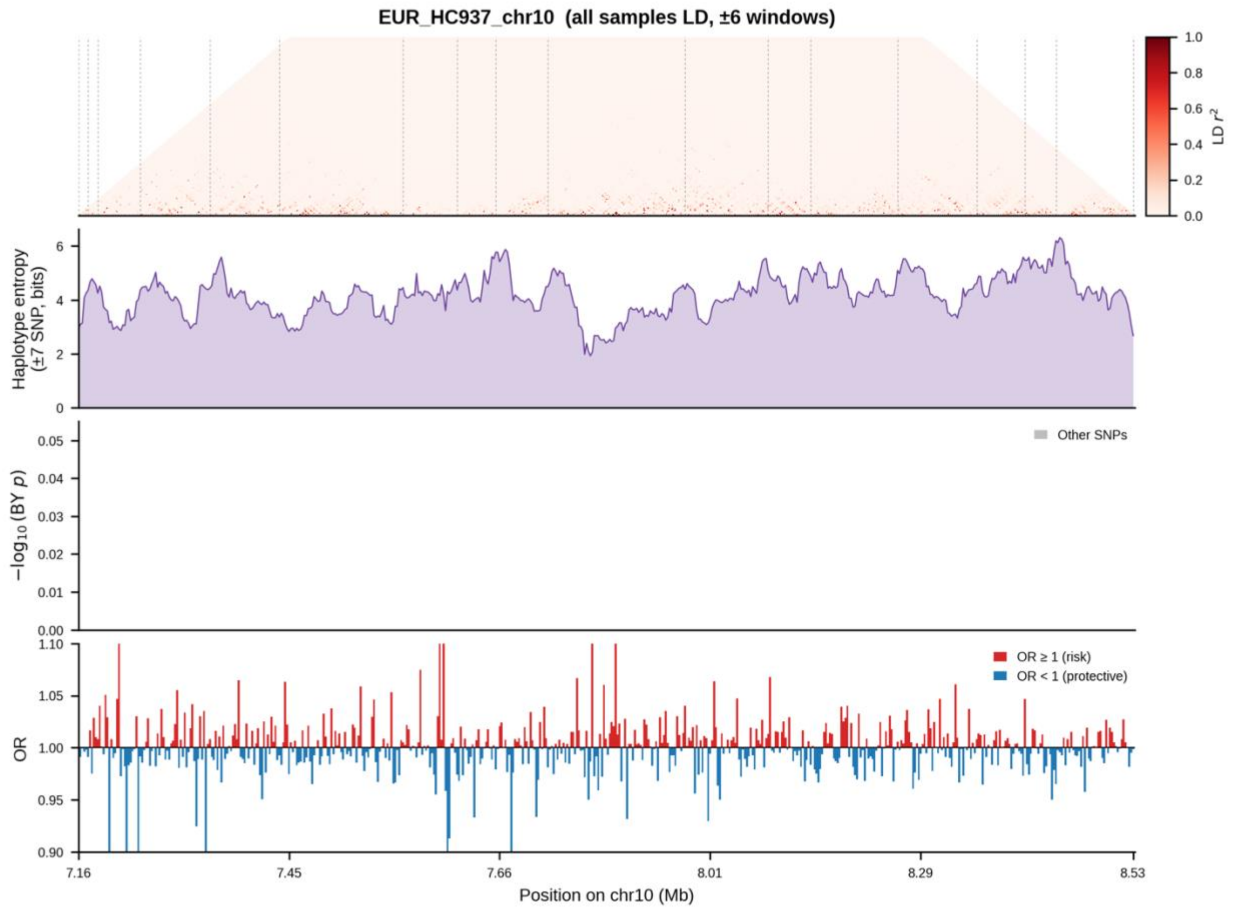


**Supplementary Figure 32**. Linkage disequilibrium, haplotype entropy, and ancestry-conditioned association signal for angina pectoris in the EUR ancestry group at the chromosome 10 locus. From top to bottom: pairwise LD ($r^2$) computed across all samples (predominantly EUR-like ancestry); local haplotype entropy (±6-SNP window, bits); BY-corrected $-\log10(p)$ values from the ancestry-associated conditional model, with the fine-mapped candidate SNP highlighted in blue; and per-SNP odds ratios, colored by direction of effect (red, OR ≥ 1; blue, OR < 1).

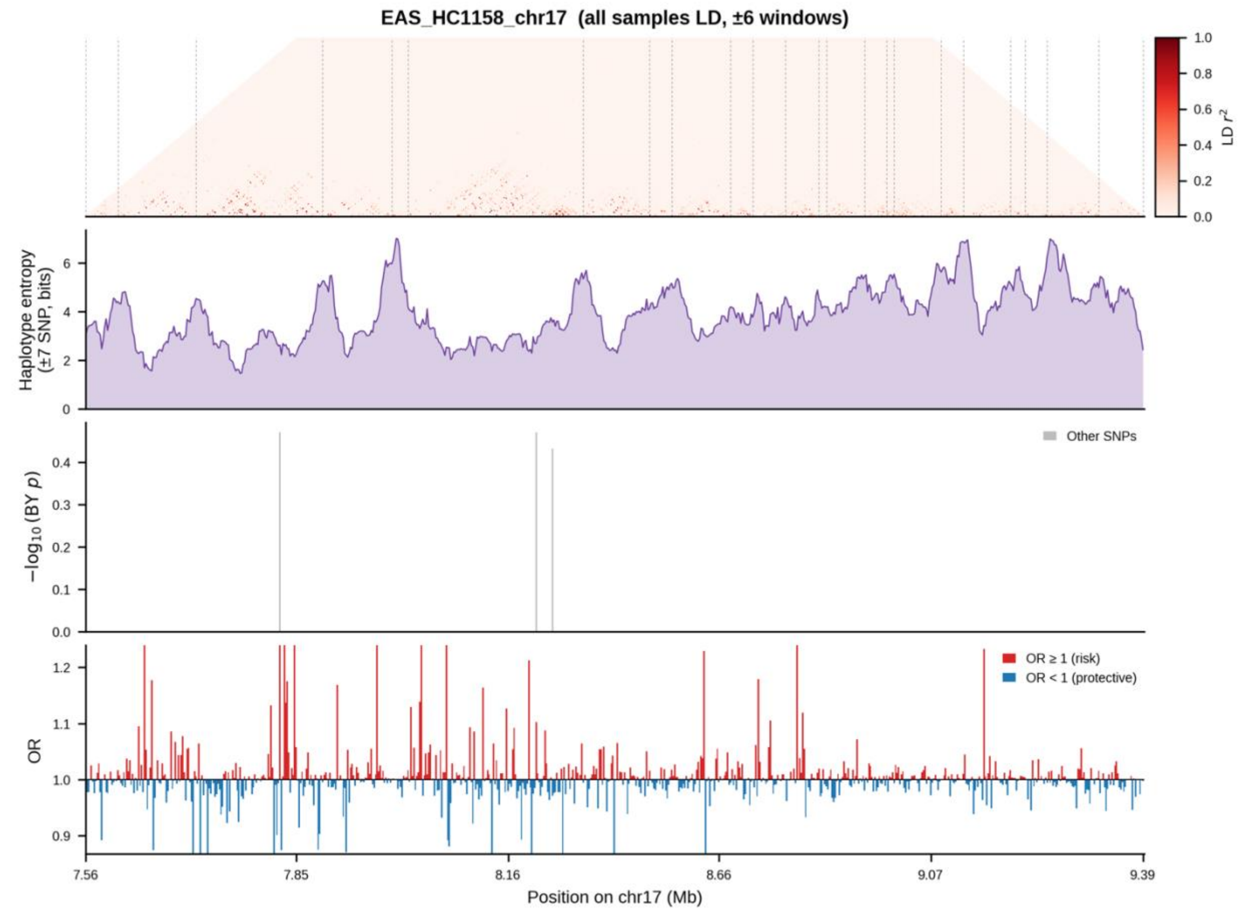


**Supplementary Figure 33**. Linkage disequilibrium, haplotype entropy, and ancestry-conditioned association signal for other dermatitis in the EAS ancestry group at the chromosome 17 locus. From top to bottom: pairwise LD (r²) computed across all samples (predominantly EUR-like ancestry); local haplotype entropy (±6-SNP window, bits); BY-corrected −log10(p) values from the ancestry-associated conditional model, with the fine-mapped candidate SNP highlighted in blue; and per-SNP odds ratios, colored by direction of effect (red, OR ≥ 1; blue, OR < 1).
